# Tailoring Human Sleep: selective alteration through Brainstem Arousal Circuit Stimulation

**DOI:** 10.1101/2023.01.18.23284688

**Authors:** Alceste Deli, Shenghong He, Benoit Duchet, Yongzhi Huang, Sean Martin, Nagaraja Sanrangmat, Timothy Denison, Huiling Tan, Vladyslav V. Vyazovskiy, Alexander Green

## Abstract

Brainstem nuclei, such as the pedunculopontine nucleus, send activating projections to cortex, modulating states of sleep, wakefulness and arousal levels. Surgical modulation of subcortical activity using deep brain stimulation (DBS) is utilised in the management of pain and movement disorders. DBS of brainstem arousal circuits in a state-dependent manner could offer an attractive alternative in severe pharmacoresistant cases of hypersomnia and in disorders of consciousness, where behavioural activation is desired.

We wanted to investigate if we can selectively induce wakefulness and/or alter sleep state through DBS of the PPN region (PPNR). To this end, we used the opportunity of implanted PPNR electrodes in stimulation-naïve patients with multiple systems atrophy. PPNR activity was recorded during both slow wave sleep (SWS) and quiet wakefulness with simultaneous cortical EEG, in order to identify differences in brainstem oscillatory patterns during different states of excitability. PPNR DBS in two gamma frequency protocols (40Hz and 100Hz) was delivered during SWS of the same sleep stage and with comparable pre-trial levels of slow wave activity.

Additionally, SHAM trials were used as a control where no stimulation was applied. We examined changes in cortical oscillatory power, changes in functional connectivity (coherence and causality) from pre- to post-stimulation and phase-locking of cortical oscillations with DBS frequencies and their sub-harmonics during stimulation. We also evaluated connectivity changes induced by DBS and corresponding differences in circuit dynamics between SWS and wakefulness.

Beta and gamma PPNR oscillatory power increased when wake was compared to sleep. We saw clear PPNR power modulation by the phase of EEG slow wave, with significant increase in gamma compared to beta power during the ‘excitable’ part of the slow-wave cycle. Gamma PPNR DBS induced transitions to wakefulness and REM, while it truncated sleep time compared to baseline. The 40Hz stimulation protocol was more efficient in reducing slow wave activity and increasing cortical beta power, compared to PPNR DBS at 100Hz. Furthermore, intrinsic cortical rhythms phase-locked with 40 Hz PPNR DBS to a significantly higher degree compared to the 100Hz protocol, with regional differences in phase-locking, suggesting a complex biological phenomenon. Finally, functional connectivity changes induced by PPNR DBS were consistent with differences in circuit dynamics between SWS and wakefulness. Overall, these results highlight the possibility of using DBS of brainstem arousal circuits to promote arousal and wakefulness, which opens new perspectives for using closed-loop approaches to modulate vigilance states in humans for therapeutic benefit.

## Introduction

Therapeutic modulation of arousal levels has broad applications but, so far, limited successes. There are currently no FDA-approved treatments for idiopathic hypersomnia, with patients relying on off-label stimulants with variable results and risks ^1^. Novel agents such as pitolisant, an inverse histamine receptor agonist, are being developed in an effort to achieve therapeutic benefit with better management of the severe side-effect profile of central simulants ^2^. Here we present evidence that electrical neuromodulation, a non-pharmacological personalised treatment, could potentially offer normalisation of sleep/wake patterns without risk of addiction ^3,4^ similar to neuromodulation for chronic pain ^5^. This may potentially be applied to sleep disorders such as severe treatment-resistant cases of narcolepsy or even rare adulthood-persistent Kleine-Levin syndrome ^6^. Outside of primary sleep disorders, decreased arousal during daytime and increases in arousal at night are also seen in disorders of consciousness (DoC), with level of impairment correlating with disease severity ^7^. A common thread of disruptions of physiological cortical activation, necessary for wake and arousal maintenance, seems to run through these conditions, as also suggested by stimulants as trial treatments for DoC ^8^.

Overall, cortical activity during higher levels of arousal is dominated by faster oscillations, contrasting with slower rhythms during sleep. Alpha and beta activity (8-30Hz) as well as gamma oscillations (>30Hz) characterize active wake ^9,10^. During deep sleep these are replaced by delta (typically 1-4 Hz) and slow oscillations (<1Hz) ^11^. However, sleep is far from a global phenomenon, with evidence of regional oscillatory patterns ^12^ influenced by levels of daytime activity ^13^, and discrete periods of cortical excitability. During slow-wave sleep (SWS), the membrane potential of neocortical and thalamocortical neurons fluctuates between depolarized (UP) and hyperpolarized (DOWN) states, reflecting the balance between network excitation and inhibition, with the emergence of cortical gamma oscillations occurring primarily during UP-states ^14,15,16^. These phenomena, as well as state heterogeneity between brain regions ^17^, may serve in evolutionarily-preserved vigilance and a level of sustained processing of environmental stimuli even during SWS ^18,19^.

A subcortical ‘sleep centre’ influencing arousal-related behaviours was first proposed in 1929 by von Economo ^20^, whose work led to the later definition of the ‘ascending reticular activating system’ modulating wakefulness and sleep by projections to cortex ^21^. This system, connecting brainstem nuclei to thalamus, hypothalamus and basal forebrain, projects to cortex through thalamic and extra-thalamic pathways modulating arousal state on both a micro and a macro scale ^22,23^. During the depolarised UP-state, ascending brainstem circuit activity seems to cluster in periods of heightened excitability. There is early evidence from rodent data that the activity of medial prefrontal cortex (mPFC), pedunculopontine nucleus (PPN) and locus coeruleus (LC) are all grouped around the UP-state of cortical slow oscillatory cycles, with PPN cholinergic neurons firing in temporal correlation with cortical gamma nested in the UP-state ^24,25^. Neuronal activity in these brainstem circuits might therefore be closely associated with oscillatory patterns present in SWS, and the modulation of these activities may further lead to the alteration of human sleep.

The PPN is also linked to the regulation of posture and movement, being part of the mesencephalic locomotor region and densely interconnected with the basal ganglia ^26^. Furthermore, basal ganglia unit activity has been shown to be temporally related to cortical UP-states, suggesting an additional indirect link between the PPN and cortical SWA^27^. These predominantly motor-related connections between the two subcortical regions have led to the PPN being surgically targeted in movement disorders (such as the Parkinsonian disorder spectrum), in both human patients ^8^ and animal models of disease^28^, thus enabling us to use this region as an ‘investigational hub’ of human *in vivo* surgical sleep depth modulation.

In this study, we use the opportunity afforded by indwelling Deep Brain Stimulation (DBS) electrodes in the PPN Region (PPNR) to investigate its role in sleep modulation. We first set out to characterize the dynamics of ascending brainstem arousal circuits contrasting sleep and wakefulness, as well as fluctuations in excitability present in SWS on a temporal micro-scale (duration of milliseconds as opposed to an entire sleep epoch). Then, we applied brainstem stimulation at gamma frequencies both at clinically utilized (40Hz), as well as at a higher frequency (100Hz) that exceeds traditionally observed physiological activity for the target nucleus, during trials of equal sleep depth, in order to assess our capability of sleep depth modulation and achieve higher levels of arousal. By showing that we can successfully decrease sleep intensity (and slow wave power as its surrogate) through short bursts of targeted neurostimulation, we highlight the potential of closed-loop, responsive systems in the treatment of pharmacoresistant hypersomnolence and DoC.

## Methods

### Ethics

This study is a substudy of a clinical trial that was conducted in accordance with the Declaration of Helsinki, approved by a research ethics committee (Preston REC 18/NW/0403) and given Health Research Authority approval. All patients provided informed written consent.

### Participants

Four male patients with a multiple system atrophy diagnosis (MSA-P, which shares features with Parkinson’s disease and falls within the same umbrella of movement disorders) and whom all had DBS leads in PPN region were recruited to this study (*Supplementary Table 1*). They were implanted with 1.5 mm-spaced St. Jude Medical Infinity directional DBS electrodes (Abbott, Plano,TX) as part of a clinical trial examining autonomic and motor function in MSA (STAG-MSA NCT03593512). We have previously described our surgical approach and technique^29^. Electrode placement was clinically confirmed after implantation by the fusion of preoperative magnetic resonance imaging (MRI) and postoperative computed tomography (CT) scans. For additional visualization of electrode positioning compared to the predicted location of important subcortical arousal-related nuclei, we reconstructed the electrode trajectories and location of different contacts using the MATLAB toolbox Lead-DBS (version 2.3.2) ^30^. The locations of the electrodes have also been reported in a previous study, which used data from the same cohort of patients ^27^. Here, we used the Harvard Ascending Arousal Network (AAN) Atlas ^21^ to co-register and normalise the contact locations into the Montreal Neurologic Institute (MNI) 152-2009b space, as well as visualise brainstem nuclei of interest in the fused images (Figure 2).

### Baseline recordings

Prior to implantation and after a night of habituation to the in-hospital environment, a polysomnographic recording was acquired (electrode placement and acquisition parameters described in ‘Supplementary Methods’). Subsequently, patients underwent electrode implantation with the electrodes remaining accessible (externalized) post-operatively, as opposed to connected to a subcutaneously implanted pulse generator (IPG). Baseline joint electroencephalogram (EEG) and local field potential (LFP) passive overnight recordings were completed after at least 72hrs of recovery from general anaesthesia, to ensure there were minimal carryover effects on slow wave activity. In addition, time of recording setup and initiation matched patient bedtime and was only undertaken once sufficient recovery had occurred and sleep-wake patterns normalized to pre-operative baseline levels.

### Stimulation Trials

Patients’ bed space was separated from the experimenter while all stimulation trials were delivered through remote programming, to avoid potentially disruptive interactions. For stimulation delivery after sleep induction in both cases (‘SHAM’ and stimulation nights), hallmarks of Non-REM Stage 2 (NREM2) sleep (K complexes/spindles and slow wave activity) were identified in a minimum of two AASM epochs preceding stimulation delivery during real-time sleep scoring (setup details in ‘Supplementary Methods’). Upon the appearance of a slow wave in the subsequent epoch, stimulation was triggered through the patient programmer from the connected ante-room/separated space, where experimental equipment was located (*Fig. 1*).

**Fig. 1.**
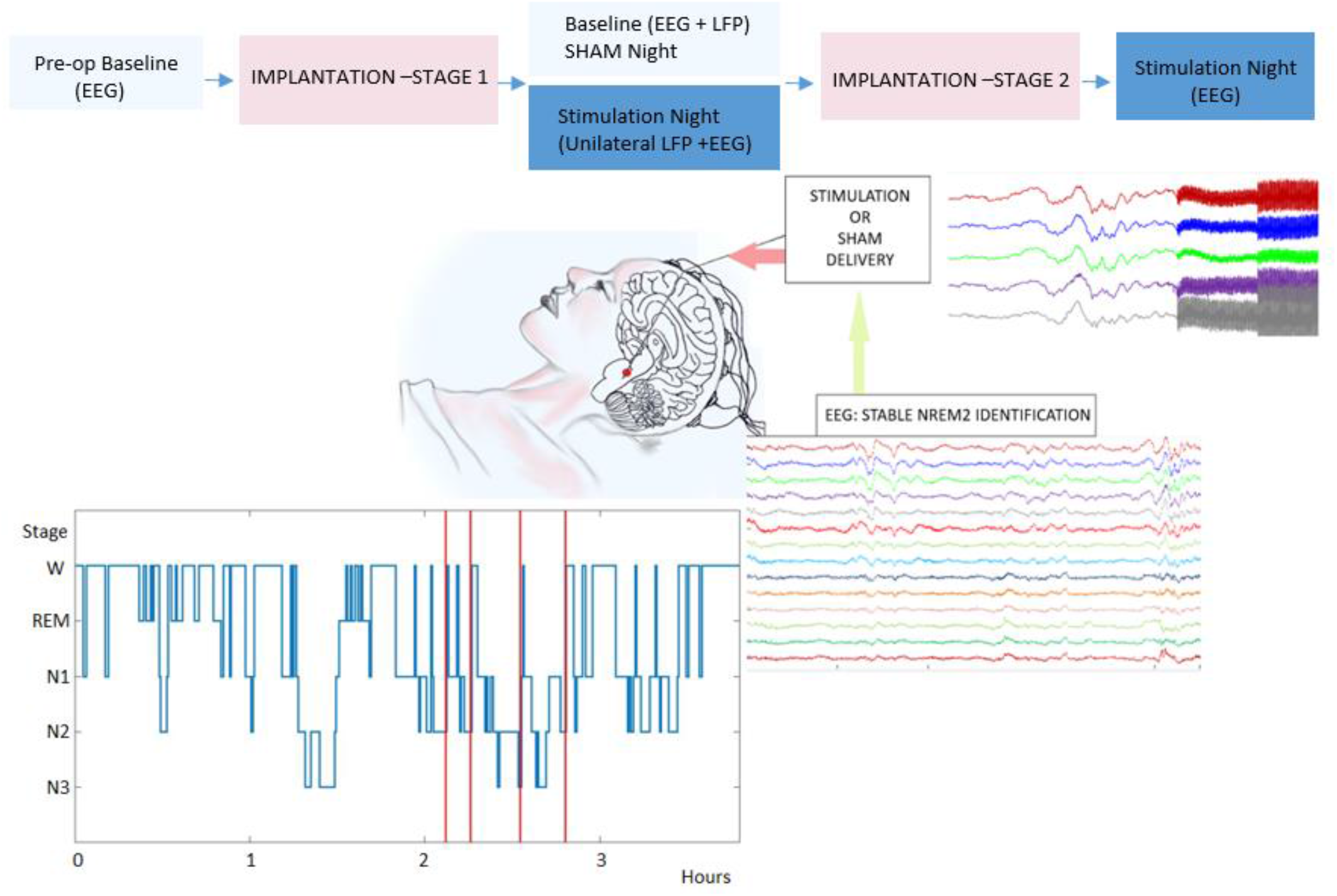
Recording blocks and visualisation schematic of an experimental trial. Top: flowchart of recording blocks (in dark/pale blue), between surgical implantation stages. Pale blue (such as in pre-operative baseline or SHAM night), denotes that no stimulation has been delivered (patients are naïve to stimulation). There is a recovery period of at least 72 hrs between each surgical stage and subsequent recording block. Bottom: Schematic of an experimental trial. Upon stable NREM2 identification on EEG (note presence of slow wave activity on a real 30-second epoch of data, bottom right) as per described criteria, stimulation or a sham trial is triggered (durations specified in text). Stimulation artefact can be visualized in the EEG data, with the amplitude ramp-up period set at a pre-thresholded value to avoid sensory disruptions (top right, where preceding slow wave activity can also be seen). A hypnogram during one stimulation night is presented (bottom-left corner), with red vertical lines denoting stimulation times during NREM2 sleep.

To fully ensure that ‘SHAM’ trials were examining circuits that were stimulation naïve, stimulation and ‘SHAM’ trials were delivered on separate nights. A night of ‘SHAM’ trials always preceded the first stimulation night, during which stimulation was delivered unilaterally through a dedicated stimulation box while the contralateral PPN activity was captured from the other DBS lead. To avoid lateralization effects, by convention we decided to stimulate the dominant hemisphere and therefore left PPN (in right-handed cases, which were 3/4) while recording from the contralateral hemisphere. Further stimulation trials were delivered on a second stimulation night after recovery from Stage 2, when both leads had been connected to the implantable pulse generator (IPG).

Pre-set rules guiding the duration of stimulation for both protocols were set in advance of randomization. When overt wakefulness occurred, the stimulation trial was terminated, while upon the observation of either stage shifts or Intra-Sleep Awakening Reactions (ISARs), the total duration of stimulation was limited to up to 3 minutes. These ISARs were defined as an abrupt, fast EEG frequency shift (including theta, alpha or frequency greater than 16 Hz with exclusion of spindles) following ten seconds of stable sleep, lasting more than ten seconds and followed by return to sleep ^31,32^. Further details on assessment of sleep fragmentation in baseline recordings can be found in ‘Supplementary Materials’. Even in the event where no effect was observed, stimulation trials to be included were limited to 5 mins of total stimulation duration. Additionally, there was a washout of at least 5 mins between trials even if sleep stage remained unaffected.

With regards to stimulation frequency, trials were randomized to two gamma stimulation protocols. One was selected on the basis that frequency would mimic endogenous PPN activity, as previously reported in the literature ^33,34^; therefore this protocol was set at 40Hz. In addition, we sought to examine the effects of ‘supra-physiological’ gamma stimulation on the circuit; therefore selected a frequency of 100Hz as potentially distinctly higher than endogenous human PPN gamma. Prior to each stimulation night, thresholding of the amplitude of stimulation parameters took place to ensure that sleep depth would not be changed by sensory/other side effects of stimulation. This was carried out by decreasing the stimulation amplitude for a defined pulse width and stimulation frequency, until an amplitude was reached where no sensory/other side effects were experienced and the patient could not identify whether the stimulation was switched on or not.

Exclusion criteria of trial data from further analyses included delivery outside of slow-wave sleep, excessive artefact presence during the post-stimulation period and/or any trials where stimulation duration exceeded standardised protocol parameters. Trial data were assessed with regards to presence of wake, ISARs and stage shifts. Finally, we only included in our analyses those trials (SHAM or stimulation) that (in addition to the above exclusion criteria) would not differ to a statistically significant degree in terms of cortical slow wave power in between the groups, therefore additionally safeguarding the fact that pre-stimulation cortical states could be considered comparable.

### Power Spectra Clustering Calculations

First, we sought to identify the LFP channels with maximal power modulations between arousal states (wake and slow-wave sleep). We therefore examined brainstem LFPs during scored epochs of quiet wakefulness and slow wave sleep (Non-Rapid Eye Movement NREM stage 3). We chose to reference our LFPs in a combination of bipolar montage configurations that preserved lead directionality while controlling for volume conduction, as has also been established practice in prior subcortical target literature ^35,36^. We selected four main frequency bands of interest to compare state-dependent signal changes: theta, 4-7 Hz; alpha, 8–12 Hz; beta, 13–29 Hz; gamma, 30–95 Hz (with a cut-off for cortical gamma activity set at 80Hz). We calculated the power spectral density (PSD) of re-referenced LFP bipolar signals using the short-time Fast Fourier transform (sFFT) with 4s of sliding-window and 3s of overlap then subsequently normalised for the noise floor of the signals during each epoch. Signals were then plotted, averaged across montage, patient and condition. In addition, the element-wise difference of such averages was calculated per patient and montage with subsequent values normalised (so that maximal and minimal values were 0 and 1 respectively), then mapped and visualised in MNI space using Lead-DBS to assess clustering of PSD changes in nuclei of interest. For subsequent connectivity analyses across each frequency of interest (FOI), LFP montages per patient that exhibited the maximal modulation across both states for the respective FOI were selected.

### Functional Connectivity Analyses

In addition to cortical power spectral changes, we sought to investigate changes in network dynamics and effective connectivity between brainstem nuclei and cortical regions –both with regards to ‘naturalistic’ differences between slow wave sleep and wakefulness as well as changes occurring after brainstem DBS. To this end, we deployed both coherence analyses (providing an estimate of coupling between the two sources) as well as causality analyses (providing more direct details about changes in direction of information flow, from brainstem to cortex).

After sleep scoring of the baseline (non-stimulation) nights, frontal channels during slow-wave sleep epochs (NREM2 and NREM3) were further visually inspected for artefact presence and overall signal quality, with artefact-free epochs selected. First, we calculated the imaginary component of the coherence between selected bipolar LFPs and frontal EEG channels. Coherence estimates are usually presented as the magnitude-squared coherence and involves its real and imaginary components. The real part, while larger than the imaginary part, is strongly affected by volume conduction effects; the imaginary part is only sensitive to synchronization of time-lagged processes ^37^. We further used Granger causality ^38^, based on a multivariate autoregression model, to assess the directionality of the connectivity (see ‘Supplementary Methods’ for further descriptions of both models).

### Brainstem Power Modulation by putative UP- and DOWN-State

The absence of direct measurements of membrane potential makes relating changes in EEG polarity during SWA difficult to directly relate to discrete UP- and DOWN-States, as these are defined in microelectrode animal literature. However, if we consider EEG as a summary of temporally synchronous cortical events, the cycle between maximum negative and positive amplitudes of EEG SWA will reflect fluctuations between hyper- and de-polarisation (therefore excitability) of the underlying cortical region. Here we therefore term this as ‘putative’ UP- and DOWN-State, as also is the case in more recent publications where this distinction between direct (microelectrode) and global (EEG) measurements of the balance between excitation and inhibition is made. ^39^

Since slow waves have been classically described as ‘traveling waves’, originating in frontal cortex before spreading to other regions ^40,41^, we mainly focused on frontal and prefrontal cortical areas for our slow-wave sleep analysis. First, we sought to identify the EEG channels with maximal slow-wave power during slow wave or Non-REM Stage 2 or 3 (NREM2/3) sleep epochs, that had been visually scored and assessed for absence of artefact. To this end, PSD of selected signals were calculated using sFFT with 4s of sliding-window and 3s of overlap and noise-floor normalisation. Then PSDs were averaged and collapsed across time, resulting in one mean value per NREMS cycle per channel within the SWA range, for selection of the main SWA frontal generator.

We then proceeded to detect cortical slow wave trains and their associated time-locked LFP activity, after which we calculated a ‘phase/power modulogram by decomposing the LFP signal in the time-frequency domain according to the putative UP/DOWN-State cycle of slow-wave activity (SWA) (‘Supplementary Methods’).

### Phase-Locking Value Calculation

The phase locking value (PLV) was used to estimate the synchronization of cortical rhythms of interest to the DBS frequency and to subharmonics of that frequency (further detail in ‘Supplementary Methods’) ^42,43^. We denoted as primary ‘bands of interest’ the alpha (8-12Hz) and beta (13-30Hz) frequency ranges. This choice was made *a priori* since these were deemed important intrinsic frequencies for excitability and sleep depth modulation, therefore the impact of DBS gamma stimulation paradigms on these should be evaluated. We focused on the gamma frequency range where dynamics between PPN and cortex significantly changed when sleep depth progressed (based on SHAM trials of this cohort). We also generated pink-noise trials in silico, to estimate the PLV that would be expected due to chance.

### Statistical analysis

Primary trial data were organised in three main groups (stimulation ‘STIM’ frequency at 40Hz, frequency at 100Hz and SHAM trials). Laterality was matched within-patient, since interhemispheric asymmetries have been documented across species ^44,45^. After individual trial data exclusion, we first ensured that sleep depth was comparable for SHAM and STIM (individual group and across both frequency groups) not only by visual scoring but also in statistical comparison of slow-wave power density across pre-stim/SHAM epochs. We used both student’s two-sample T-Test (individual stim group, SHAM) per each frequency band within group and a two-way ANOVA across groups, while Bonferroni correction was deployed for multiple comparisons. Once this homogeneity was established, data from the stimulation and SHAM cohorts were further analysed (*Supplementary Statistics*).

## Results

During baseline recordings, significant differences in oscillatory activity were seen between wake vs sleep (macro-state, Figure 3A) and putative UP/DOWN states, as well as associated underlying circuit functional connectivity changes. In addition, we were able to significantly alter sleep stage and brainstem-cortical connectivity through selective PPNR stimulation.

**Figure 2.**
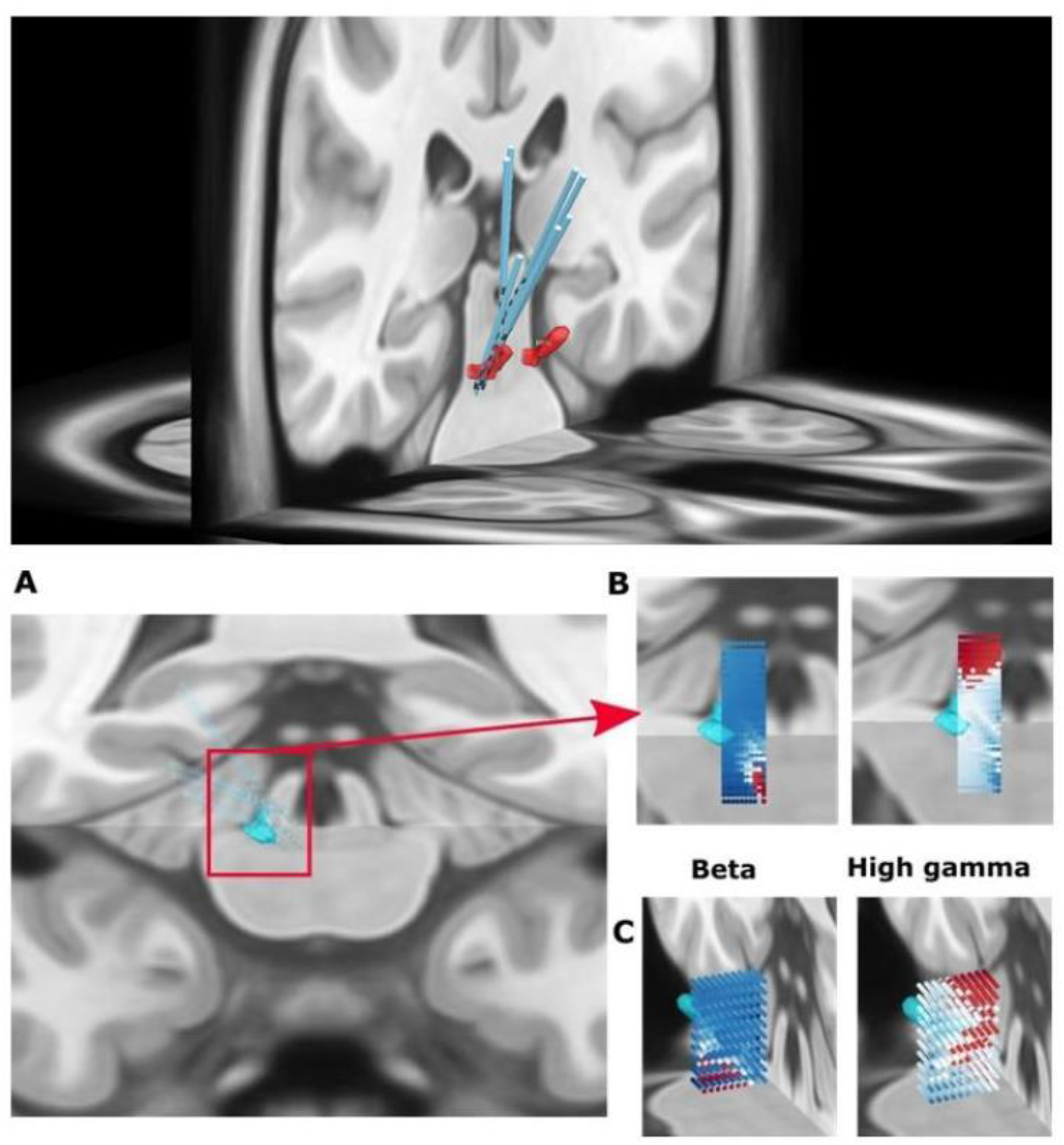
Distribution of Power Modulation between Wake and Slow-Wave Sleep (SWS) for Brainstem Local Field Potentials (LFPs). Coronal section (top) with all mirrored leads positioned for orientation purposes (rostro-caudal and anterior-posterior), utilizing localization information from MRI (pre-operative) with post-operative CT scans from all patients. The relevant power differentials are then plotted on the location of each lead contact, in MNI space, with a colour gradient reflecting degree of modulation (red for higher differential values versus blue for lower). More anterior and caudal origins of beta modulations are noted. High gamma activity outside but adjacent to the PPN nucleus on the other hand, was mainly located rostrally but towards more posterior regions. The PPN region is plotted in MNI space using the Harvard Ascending Arousal Network (AAN) and visualised here in pale blue. Figures were all generated using Lead-DBS.

### Gamma and Beta PPNR Oscillations are Maximally Modulated by Fluctuations in Excitability across different temporal scales

Frequency-specific oscillatory activity in the PPN was modulated by the phase of cortical slow waves. Maximal differences in power modulation were observed for beta/low gamma and high gamma when assessing temporally macroscopic differences in excitability, by comparing SWS and quiet wakefulness. Specifically, beta/low gamma (19-34Hz, p=0.0311) as well as higher gamma (77-95Hz, p=0.0195), increased when wake was compared to sleep (*Figure 3B, left panel*). This change at the ‘macro’ level (sleep vs wake) was also visualised by plotting the linear power spectra across re-referenced pairs per patient hemisphere (*Figure 3A)*. Despite wake-related changes in the alpha (8-12Hz) frequency band when brainstem LFP power spectra were visualised, this effect did not reach statistical significance based on the power modulogram (*see Supplementary Methods)*. Additionally, it seems that many such contacts with higher beta and gamma modulation clustered around the PPN area when power modulation was plotted against the AAN atlas in MNI space (*Figure 3C)*. There was a distinct distribution with higher gamma modulation rostro-caudally, when LFP power spectral modulation was assessed after plotting on an anatomical axis, suggesting underlying differences in state-dependant modulation of brainstem neuronal population activity *(Figure 2*).

**Figure 3:**
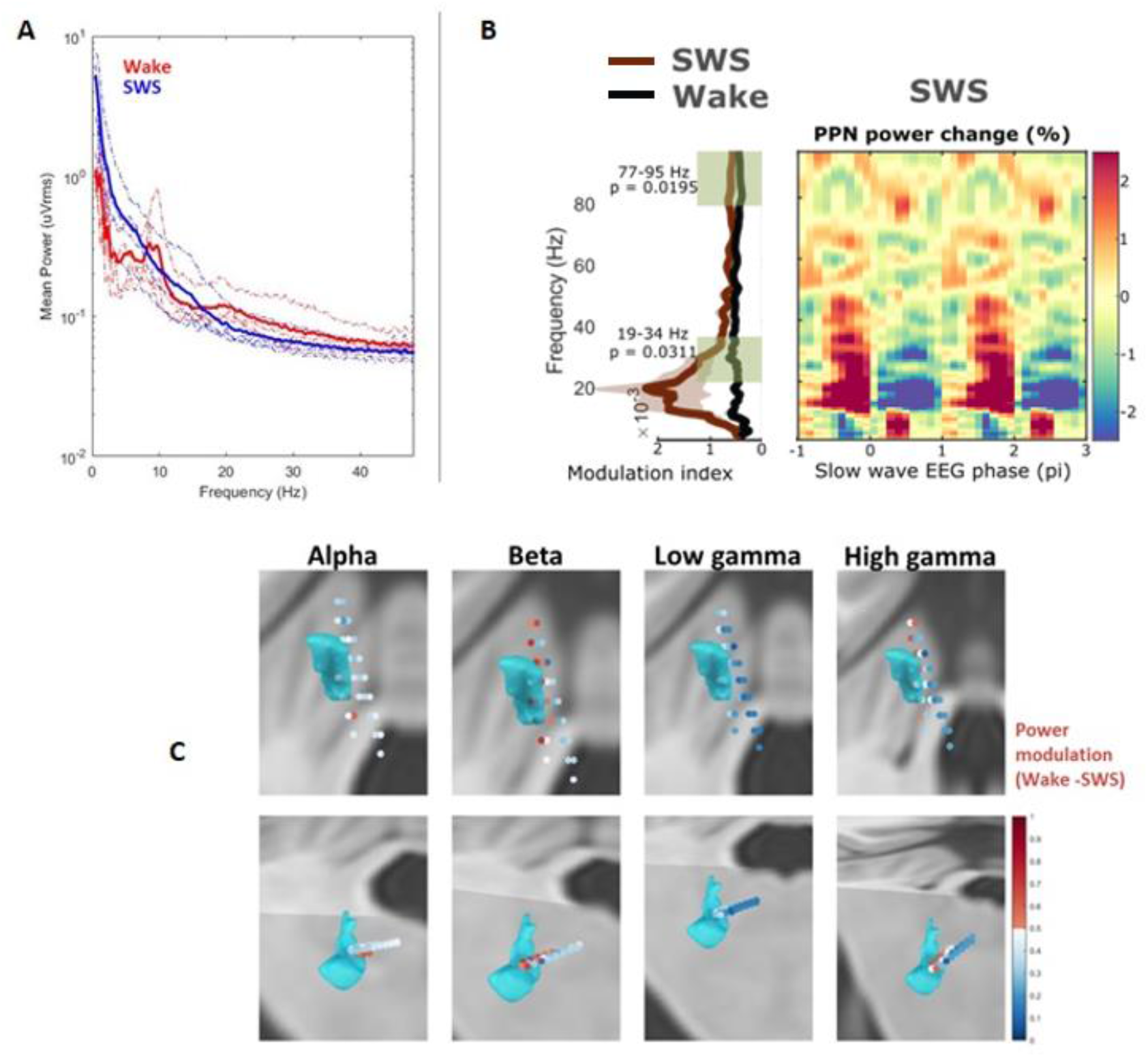
Power Modulation between Wake and Slow-Wave Sleep (SWS) for Brainstem (PPN Area) Local Field Potentials (LFPs). A. Linear plots of average LFP power across patients for two macro-states (wake: red, SWS: blue). Note the beta and alpha peak during wake-state. B. Statistical differences in modulation index across the power spectra (left panel) and modulation index variations during SWS (with slow wave phase). Statistical analyses across eight hemispheres (and respective LFP channels). C. LFP power modulation across lead contacts plotted in LeadDBS. Note high modulation of beta, high gamma and their clustering around the PPN area (light blue shape, methodology and rationale as per Figure 2 legend and ‘Methods’ section).

On a temporal ‘micro’ scale (msec as opposed to multiple sleep epochs), we also saw clear PPN power modulation by the phase of EEG slow wave during sleep (Figure 3B and Figure 4A, depicting two putative UP/DOWN State cycles). When we made comparisons with power modulations during equivalent 2pi cycles in quiet wakefulness data, where this periodic fluctuation between excitation and inhibition is not present this effect was not observed (*Supplementary Figure 4*). Beta and high gamma power were anti-phased with regards to the slow wave phase (*Figure 4B)*. Based on the average of all re-referenced LFP signals, high gamma power started building up during the threshold-crossing and was maximal during the peak of the UP-state (p=0.001) (*Figure 4B)*. This resulted in a statistically significant increase in gamma compared to beta power (p=0.0078, 10000 times permutation test) during the ‘excitable’ part of the slow-wave cycle (transition to maximal positive peak of the EEG slow wave/putative UP-State, Figure 4B). Changes in alpha modulation did not reach significance for the total average of all re-referenced LFP signals.

**Figure 4:**
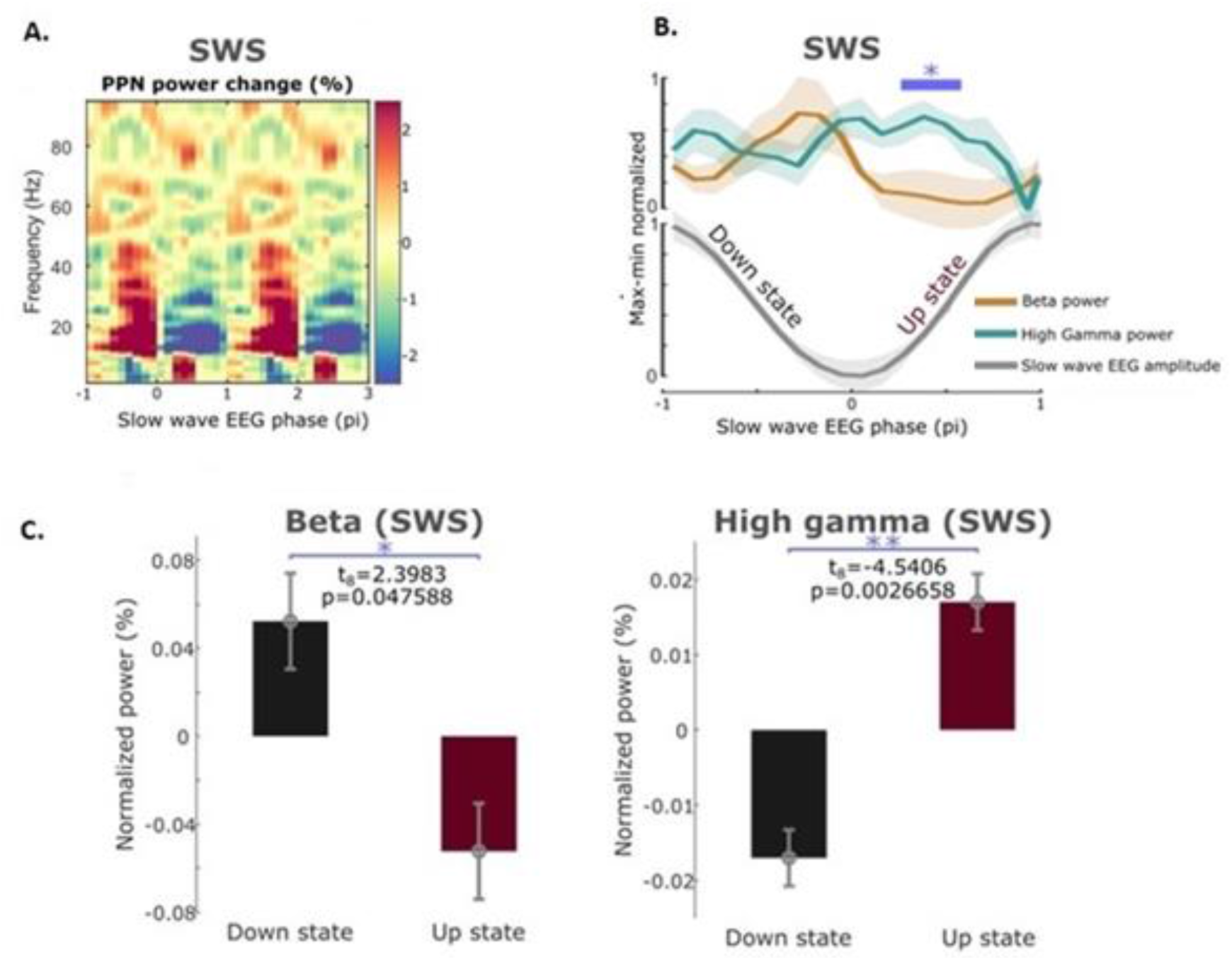
Brainstem LFP power changes with regards to putative UP/DOWN State (Slow Wave phase). A. Power modulogram during slow wave sleep (slow wave snippets, as further described in Supplementary Methods). Maximum power modulation (%percentage compared to average normalised power-in-band) clusters around the first (negative) phase of the slow oscillation cycle (here termed as a reflection of cumulative hyperpolarisation/DOWN-State) for beta and around the second (positive) phase of the slow oscillation cycle (cumulative depolarisation/UP-State). B. Statistical differences in modulation index differences (maximum minus minimum) for beta and high gamma, across slow-wave phase. Note the significant difference between the power in these two frequencies during the positive phase of the EEG slow wave (p=0.0078). C. Further comparisons of beta and high gamma frequency (for signals with higher beta and lower high gamma frequency during the putative DOWN-state), providing additional evidence of anti-phasing for these two oscillations.

### Intermittent PPNR Gamma Stimulation during SWS changes Sleep Architecture

During stimulation experimental nights, despite the stimulation being delivered below sensory thresholds, trials with PPNR stimulation frequency of both 40 Hz and 100 Hz resulted in an increased frequency of stage shifts and sleep disruption, compared to baseline ‘OFF’ nights. Overall sleep time was truncated since spontaneous arousal occurred earlier than during the baseline night, with patients unable to resume their usual sleep schedule (*sample case Fig 5*).

**Figure 5.**
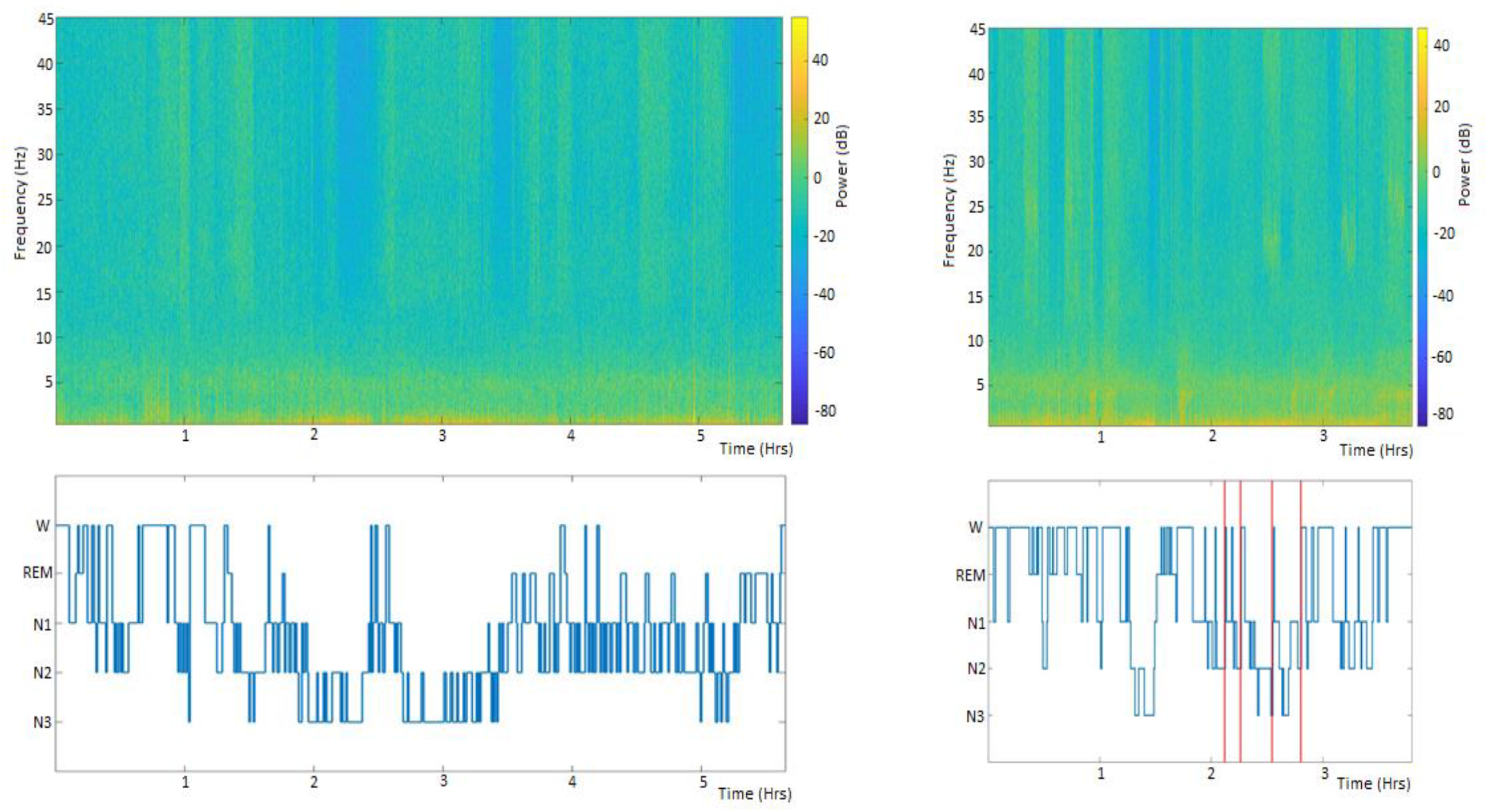
Sleep stage transitions and wakefulness induction through bilateral PPN stimulation. Left: baseline sleep in stimulation-naïve patient. Note the prolonged period of deep slow-wave sleep (NREMS2 and 3) that occurs between 2 and 3hrs 30 mins. This was targeted with stimulation in the bilateral stimulation night (right). Right: fragmented sleep with multiple micro-arousals during SWS and limited time with slow wave activity during the stimulation night. Please also note that time until awakening differs (spontaneous in stimulation night, with behavioural hyperarousal in this patient, compared to previously timed alarm to end recording at 6hrs during the baseline night).

We ensured that we were comparing stimulation trials to SHAM trials with the same pre-stimulation slow wave characteristics, in addition to visual scoring of the preceding sleep epochs, for optimal stage comparability. There was no statistically significant difference in terms of slow wave activity (SWA) power between the pre-stimulation and pre-SHAM period (p =1, CI −0.1630 to 0.1237), Across all comparable stimulation trials (*see Supplementary Results*), wake-associated oscillations increased and slow wave activity mainly decreased even when wakefulness did not immediately ensue (also reflected in trial spectrograms, *Figure 6I*). Overall, arousal-related events (both wake epochs and ISARs) increased by 33.83%. Transitions to REM stage were increased by 16.91% following both stimulation protocols, making this the most likely transition outcome (*Table 1*). We proceeded to formally analyse changes in slow wave, alpha, beta and gamma power (two-way ANOVA with Bonferroni correction for multiple comparisons).

**Figure 6.**
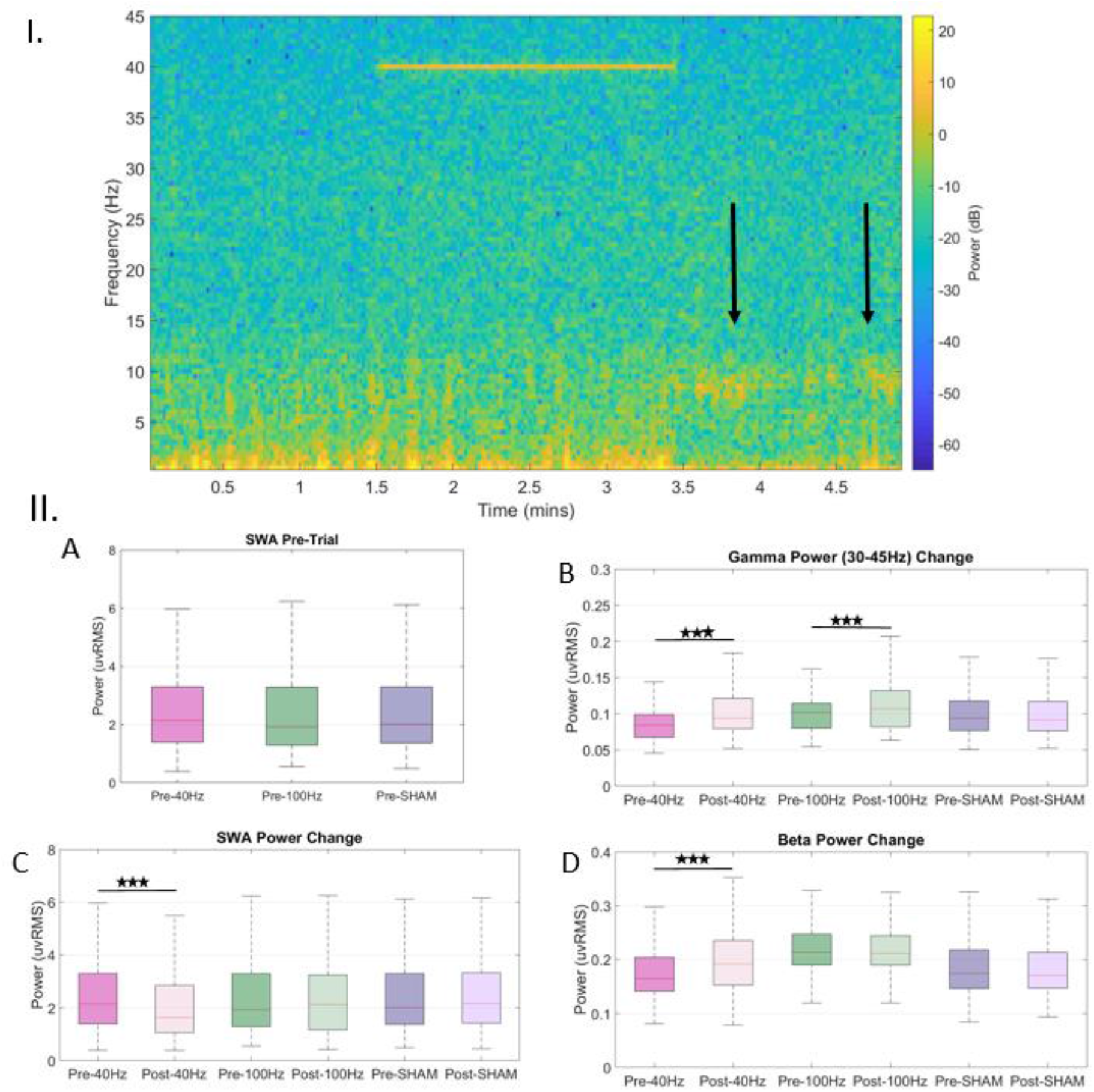
I. <u>Sample trial spectrogram (40 Hz unilateral stimulation)</u>. Arrows: Increase in alpha power and decrease in slow wave power is evident post-stimulation (arrows), while the patient transitioned to an epoch of full wakefulness following this trial. Stimulation artefact is clear (at 40Hz). II. <u>Differences in oscillatory power changes between the protocols</u>. A. Pre-trial Slow Wave Activity (SWA) power does not differ significantly between trial groups (40Hz, 100Hz, SHAM), suggesting comparable sleep depth. B. Both 40Hz and 100Hz stimulation trials lead to a significant increase in cortical (EEG) gamma activity (here shown in the 30-45Hz range). C. Only 40Hz stimulation leads to statistically significant decreases in SWA power, compared to pre-stimulation levels. D. Only 40Hz stimulation leads to statistically significant increases in beta power, compared to pre-stimulation levels. Alpha power changes (significant increases after both protocols) not depicted in this graph set; described in section text.

**Table 1.** Stage transitions following stimulation and SHAM trials. Outcome was evaluated based on the first three AASM epochs following trial duration. Should there be further stage fluctuation post-trial, the first epoch was taken into account (for instance a REM/REM/NREM would be accounted for as REM trial outcome).

| **SHAM trials** |  |
| --- | --- |
| **No stage transition** | 50% |
| **REM** | 6.25% |
| **Wake** | 6.25% |
| **ISARs** | 18.75% |
| **Stage Depth progression (NREM3)** | 12.5% |
| **STIM trials** |  |
| **No stage transition** | 23.53% |
| **REM** | 29.41% |
| **Wake** | 23.53% |
| **ISARs** | 17.65% |
| **Stage Depth progression (NREM3)** | 0% |

Both stimulation protocols (40Hz and 100Hz trials) were characterised by significantly increased high and low gamma oscillations, compared to SHAM trials (p<0.0001) as well as alpha oscillations (p<0.005) (*Supplementary Results)* (*Figure 5IIB)*. However, the two protocols showed differences based on the magnitude of the stimulation effects on slow-wave activity (SWA) and beta power. The 40Hz (‘physiological’) stimulation protocol led to a significant decrease of SWA post-stimulation (p <0.0001, CI 0.14397 to 0.3052) and an increase in beta oscillatory power post-stimulation (p<0.0001, CI −0.0314 to −0.0153). In the case of the 100Hz matched stimulation trials, these effects did not survive the Bonferroni correction for multiple comparisons (p =1, CI −0.1854 to 0.3555 and p =1, CI −0.0081 to 0.0121 respectively). In the case of the SHAM group, there was no significant change in slow wave power when the pre-SHAM and post-SHAM periods were compared (p =1, CI −0.08722 to 0.0414) (*Figure 6IIC, D)*.

We subsequently compared all pooled matched stimulation trials (n=22) to an equivalent number of SHAM trials delivered at comparable sleep stage (NREM2) and SWA power. SWA significantly decreased post-stimulation (p <0.0001, CI 0.2841 to 0.5333) but not post-SHAM trial delivery (p =1, CI −0.1791 to 0.1435). Additionally, alpha power increased during the post-stimulation period (p <0.0001, CI −0.057 to −0.0306), as well as beta (p<0.0001, CI −0.0185 to −0.0097) and so did low (p<0.0001, CI −0.0168 to −0.011) and high gamma cortical oscillatory power (p<0.0001, CI −0.0163 to −0.0107). However there was no change in those frequency ranges during the post-SHAM period (p>0.1 in all cases).

### Brainstem-Cortical Connectivity changes Post-PPNR Stimulation resemble Wakefulness and fragmented Sleep

State-dependant modulation of imaginary coherence (ICoh) values between EEG channels with maximal SWA power and PPNR revealed a significant increase in gamma ICoh during wake, with the cluster of maximal importance centred around 68Hz (range 63-74Hz, p= 0.025 t= −3.1533). No statistically significant changes occurred within the low gamma or beta frequency range. Subsequently, ICoh values for interactions with all prefrontal and frontal channels were calculated during both SWS and wake. Gamma ICoh between cortex and brainstem was significantly higher during wake. This occurred again within the high gamma frequency range (61-64Hz p= 0.0060 t= −4.6370). No statistically significant changes occurred for ICoh within the low gamma (30-55Hz) frequency range. Inversely, beta ICoh between frontal cortex and brainstem increased during sleep compared to wake for this collective channel group (15-16Hz p= 0.001, t= 2.6439).

When the most significant cluster was examined across the entire frequency range for all prefrontal and frontal channels, it was in the high gamma range (60-63Hz, p= 0.005, t=-4.6370). This change closely resembled the main post-stimulation cluster, when clinically used stimulation was applied and compared to prior SWS (59-67Hz, p= 0.004, t= −4.6007) (*Figure 7A*). We focused on the effects of the clinically used stimulation frequency on bottom-up circuit dynamics, since this also showed the greatest effect on SWA reduction in matched and controlled trials). When changes in each frequency band were examined separately, a significant increase in coherence in the beta frequency range also occurred after stimulation, present close to the half-harmonic of the contralateral stimulation for these channels (14-19Hz, p =0.024 and t= −2.7488). In all pooled ‘SHAM’ epochs, gamma coherence between frontal cortex and brainstem decreased, as evidenced for both lower (39-44Hz, p =0.014 and t= 2.9957) and higher (75-79Hz, p =0.016 and t= 2.9818) ends of that gamma spectrum (*Figure 7B and Supplementary Figure 5*).

**Figure 7.**
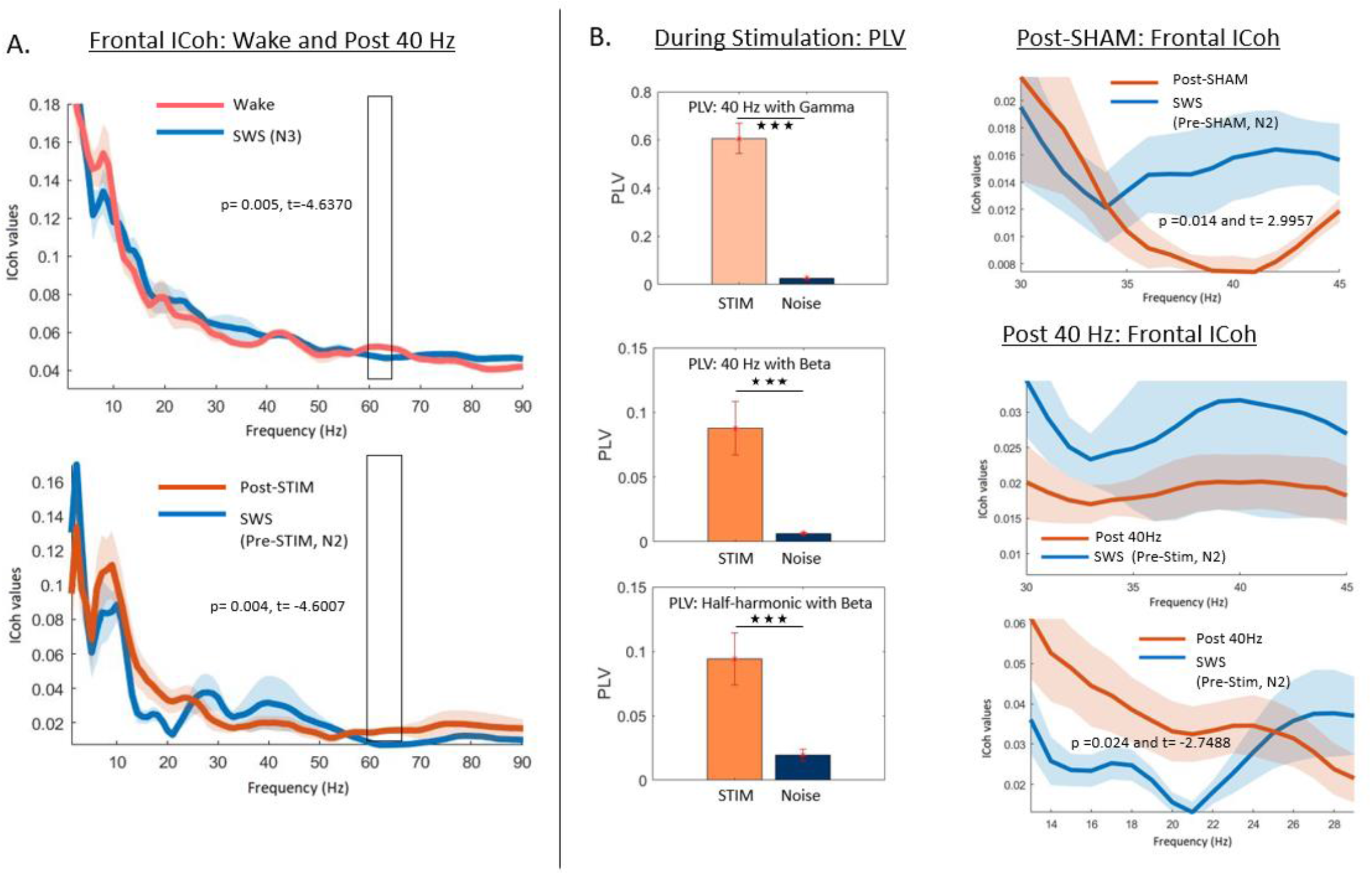
A. <u>Comparisons between imaginary coherence values (ICoh) between PPNR and frontal EEG channels in wakefulness, SWS and after 40 Hz unilateral stimulation</u>. Frequencies with maximal ICoh values show overlap between wakefulness and after stimulation (further details in main text). B. <u>Phase-Locking Values (PLV) between stimulation at 40Hz and endogenous rhythms (beta and gamma oscillations in frontal EEG channels) (left column)</u>. At stimulation frequency, there is significantly higher PLV with endogenous beta and gamma compared to pink noise. Additionally, the half-harmonic of the stimulation shows significant phase-locking with endogenous beta. <u>Imaginary coherence (ICoh) changes after SHAM trials and 40 Hz stimulation (right column)</u>. Frequency band for beta and gamma surrounding the stimulation frequency.

There was no significant difference in wake GC values between any of our patients, across all examined frequency ranges (p>0.1 in all cases). However, in all cases, bottom-up information flow (from brainstem to cortex) increased significantly across the beta and gamma ranges during SWS (p<0.00001 in all cases, after Bonferroni correction). This suggested the presence of a common process active in all patients and was correlated to degree of sleep fragmentation (*see Supplementary Results*). Post-stimulation periods were characterized by an increased gamma information flow to cortex compared to pre-stimulation trials, with frequencies of increases both in the lower and high gamma range. For the low gamma range, permutation clusters centred at 43Hz and 45Hz, with p =0.014 t= −2.2220 and p =0.002 t= −2.0654 accordingly, while at the high gamma range, permutation clusters centered around 65Hz (p =0.005 and t= −2.5072), 75Hz (p =0.01 and t= −2.2573) and 81Hz (p =0.012 and t= −2.1524). There was no statistically significant change in information flow in the ‘SHAM’ stimulation group, where sleep depth remained the same for more trials compared to stimulation trials (26.47%), or in some instances even increased.

In order to assess globally present, as opposed to regional (frontal or occipital) changes, we additionally compared and visualized differences in Granger causality outcomes between SHAM trials and stimulation trials (as opposed to within-group comparisons), based on the difference between pre-stimulation and post-stimulation values in each case (*Supplementary Figure 6*). When stimulation values were directly compared with SHAM, permutation clusters of the most significant differences centered around the half-harmonic of the stimulation frequency (21Hz p =0.001 and t= −2.26075, 23Hz p<0.001 and t= −2.2803, 25Hz p =0.001 and t= −2.21727) as well as higher gamma values (65Hz p =0.003 and t= −2.82713, 67Hz p =0.003 and t= −2.82713, 69Hz p =0.009 and t= −2.45279, 73Hz p =0.21 and t= −2.29470, 81Hz p =0.005 and t= −2.61225).

### Cortical Beta and Gamma Oscillations Significantly Phase-Lock to Brainstem Stimulation Frequency

We subsequently examined phase-locking between the DBS stimulation frequency of either protocol and spontaneous cortical rhythms, as this would provide further proof of synchronization at key frequencies during the stimulation period, initiating the differences in activity that were observed when the stimulation has stopped. We analysed trials from three patients, pooling all 21 frontal and pre-frontal dipoles resulting from these trials while analyzing all 18 occipital dipoles as a separate group. We only used trials of the same laterality within each subject (not combining bilateral and unilateral trials due to differences in amplitude thresholding, which may potentially impact results). We proceeded to examine regional differences in phase-locking, suggesting a level of variance per cortical brain area in terms of phase/oscillatory synchrony, which would also be in keeping with a complex biological phenomenon (*Supplementary Results)*.

We initially focused on the clinically used, ‘physiological’ stimulation frequency (40Hz). Frontally and occipitally, endogenous beta and gamma significantly phase-locked to stimulation frequency (p<0.0001), while cortical beta also significantly phase-locked to the frequency’s half-harmonic (p<0.0001, Figure 7B, see *Supplementary Results for full statistical results and mean PLV values)*. Occipitally, endogenous cortical alpha phase-locked to the quarter-harmonic of the stimulation frequency significantly higher pink noise (p =0.0098, zval=-2.582), a finding that was absent from frontal regions (p =0.8013, zval=-0.2517). A similar finding occurred in the case of 100Hz stimulation with occipital phase-locking of endogenous alpha to a significant degree, compared to its association with pink noise control signals (p =0.0106, zval=-2.5545) that did not occur frontally (p =0.2018, zval=1.2766). Here however, homologue endogenous (beta) oscillations did not phase-lock to the quarter-harmonic of the stimulation frequency neither frontally (p =0.4009, zval=0.8401) nor occipitally (p =0.3825, zval=0.8733). Gamma and beta oscillations significantly phase-locked with stimulation at 100Hz on both sites (p<0.001, *Supplementary Results)*.

We further proceeded to examine whether phase-locking of endogenous cortical rhythms was more significant for one stimulation protocol versus the other, for pooled trials from all patients. With regards to alpha oscillations, both protocols showed the same degree of phase-locking of the frequency of interest frontally (no statistically significant difference, p= 0.9403, zval= 0.0749). However, occipitally, phase synchrony between the 40Hz stimulation protocol and endogenous alpha oscillations was significantly higher compared to that of the 100Hz protocol (p= 0.0053 zval= 2.7862). With regards to cortical beta, endogenous oscillations showed higher phase-locking with 40Hz stimulation both frontally (p= 0.0128, zval= 2.4893) and occipitally (p< 0.0001, zval=3.5430), while gamma at the frequencies of interest also significantly phase-locked with this protocol (p<0.0001). This higher degree of phase-locking survived further analyses with a Bonferroni correction for multiple comparisons.(two-way ANOVA, *see Supplementary Results*)

## Discussion

We successfully altered sleep depth, quadrupling the probability of transition to wake through PPNR gamma stimulation. This effect is also highlighted by the fact that post-stimulation PPNR-cortical dynamics resemble those changes observed when wakefulness is compared to SWS as well as during sleep fragmentation, where sleep depth is not maintained. Gamma stimulation of the PPNR successfully enhanced cortical gamma activity, while the additional fact that phase-locking of endogenous rhythms to stimulation was regionally variable suggests a complex biological phenomenon of oscillatory synchrony. The comparative strength of the 40Hz stimulation protocol in reducing SWA and enhancing cortical beta may lie in its greater ability to phase-lock of cortical beta with its half-harmonic in addition to cortical gamma, therefore enhancing a broader range of wake-related oscillations compared to the 100 Hz protocol. One could however argue that the lower amplitudes necessitated by the lack of patient tolerance of supra-physiological gamma might have also played a role in this.

Gamma PPNR stimulation however also increased the probability of REM transitions, to an equal degree as transition to an awake state. This result is consistent with *in vivo* evidence from other species, as well as the dominant hypothesis that the PPN and LC form part of a ‘REM ON’ switch ^46^. Initial evidence from trials of electrical stimulation applied to the PPNR in freely-moving cats, showed increased REM sleep time as a result ^47^. This was later followed by selective microinjections of glutamate in areas with cholinergic PPN neurons, where it was observed that at lower concentrations REM sleep would be induced, while wakefulness ensued upon higher levels of imposed excitation ^48^. This early finding could explain the dichotomy in our outcomes, potentially depending on the number of cholinergic neurons recruited. REM induction and an increase in cortical gamma was also observed in later optogenetic stimulation of cholinergic populations ^49,50^.

With regards to humans, one longitudinal case report mentioned an increase in total REM sleep time after longitudinal PPNR stimulation ^51^, while a study in five patients during ON/OFF unilateral PPNR stimulation periods reported selective doubling of REM sleep time ^52^. The stimulation parameters in these studies differed from ours; consisting of empirically-determined frequencies, different from ours (25Hz in the first study and 5Hz, 30Hz and 70Hz in different patients of the second study, respectively). This fact, as well as lead localization differences could explain the REM-selectivity, especially in the latter study. As also highlighted by the authors, DBS electrical field could affect LC afferents contributing to a REM-modulating effect (especially in cases of deeper implantations). Another group explored higher stimulation frequencies, however during the day only, reporting that while 25Hz had a wake-promoting effect, PPNR stimulation at 75Hz caused episodes of ‘irresistible sleep’ ^53^. This result, unique in the literature, was partially explained as a DBS-mediated lesional effect. However there is evidence that a lesional effect may actually have the opposite result; in a study of 35 Wistar rats where the PPN was lesioned, sleep became disorganised with higher probabilities of Wake/REM transitions, without any acute ‘irresistible sleep’ effects ^54^. It is entirely plausible however that the region implanted in this human study varied significantly from its rodent homologue and/or what other centres (including ours) currently use as a target for PPNR implants.

In our sample, we noted that there was differential state-dependent oscillatory modulation across the lead, with rostral portions of the implanted region showing greater modulation of gamma frequencies while in caudal and anterior portions beta modulation predominated. The fact that the PPN is a heterogeneous structure comprised of both cholinergic and non-cholinergic populations could translate to some differences in neuronal population dynamics, with published evidence in other species. In an *in vivo* study of urethane-anaesthetised rats, where SWA and UP/DOWN states are present mimicking natural sleep, PPN neuronal firing was primarily coupled to the peak of the UP-State of the cortical slow oscillation in one cholinergic population (44.2%), however in another, main population firing was modulated by the DOWN-State (11.1%) ^55^. In our human cohort, we found that (caudally-predominant) beta oscillations clustered during the DOWN-state while rostral gamma activity was more prominent during UP-states.

One could theorise that two processes/population samples have joint roles in promoting PPNR-mediated excitation: one first ‘driving’ transition to a higher excitatory drive (with beta accumulation during DOWN-state). Subsequently, after a threshold is crossed, a second population is activated and permitted to fire during the ‘active’ slow wave phase, whose increased activity maintains it and coincides with the peak of the cortical UP-state (gamma burst during UP-state). The presence of discrete populations with the capability of tuning arousal has also been investigated in brainstem nuclei such as the locus coeruleus, where a GABA-ergic sub-population directly inhibits a local activating noradrenergic sub-population based on environmental cues, tuning the nucleus’ response ^56^.

The fact that higher information flow (as quantified by GC) between cortex and PPN is present in cases of greater sleep fragmentation in the beta and gamma range could reflect the underlying hyperactivity of this dual mechanism in the context of circuit neurodegeneration. Insomnia as a feature in MSA, attributed to both primary neuropathology as well as secondary factors (anxiety at bedtime, sleep fragmentation secondary to freezing, nocturia or presence of periodic limb movements) ^57,58^. Supporting our finding of PPNR hyperdrive associated with sleep fragmentation, in a relatively recent study of primates with acute Parkinsonism, beta coherence between basal ganglia and cortex increased prior to spontaneous awakenings compared to healthy controls. Comparatively higher subcortical beta power during sleep in the affected animals was correlated with decreased NREM sleep and more frequent episodes of fragmentation ^59^.

Our study’s pathological substrate, as well as the limited number of patients, could be considered a significant limiting factor to the generalization of our findings. However, there is a great degree of concordance with studies across time and species, utilizing various stimulation modalities. We are therefore hoping to lay a roadmap towards responsive, closed-loop (CL) DBS protocols modulating sleep and wakefulness. Such protocols could be tailored according to either macro or micro-scale of circuit excitability –although direct, spike-based approaches ^60^ may not be currently feasible with human implants. In the vein of our two-population hypothesis, we would expect a protocol, delivering stimulation that enhances endogenous PPNR beta during the DOWN-state and gamma during the transition to the cortical UP-state, to be the most efficient at promoting wakefulness through maximal cortical activation. Such a tailored protocol could be used to avoid sleep bouts in cases of hypersomnia.

Additionally, sustained enhancement of PPNR gamma activity during daytime could boost alertness and prevent excessive daytime sleepiness. Especially when advanced neurodegeneration impacts the ability of cortical gamma entrainment through non-invasive means ^61^, PPNR DBS at gamma frequencies could be an attractive solution for modulation of alertness and cognitive performance. However, in individual conditions, further tailoring of settings according to endogenous activity would be crucial to success and avoidance of side-effects. For instance, REM enhancement in narcolepsy could be a problematic point, potentially further exacerbating a NREM-to-REM transition dysregulation at the core of the disorder ^62^. Another question is whether habituation to the effects of PPNR on arousal would occur in chronic trials and as disease progression ensues.

Longitudinal trials of CL protocols are required to safely answer this question in a variety of conditions. Furthermore, the inverse question (enhancing sleep depth through brainstem arousal circuit stimulation) is an additional area under investigation –either through suppression of pathological hyperactivation or through entrainment of cortical SWA. In addition to frequency considerations, the stimulation mode (burst as opposed to tonic) is another factor to consider in this context, given the intermittent patterns of activity that these circuits physiologically exhibit. Novel devices capable of sensing endogenous activity and reliably delivering CL stimulation as a response to state changes, are an additional pre-requisite for these new treatments and an area of active collaborative research for our group ^63^.

## Supporting information

Supplementary Meterial

## Data Availability

The authors will consider requests to access the data that support the findings of this study in a trusted research environment.
Contact: Alceste Deli

## Data Availability

The authors will consider requests to access the data that support the findings of this study in a trusted research environment.

Contact: Alceste Deli

