## Supplementary Meterial for "Tailoring Human Sleep: selective alteration through Brainstem Arousal Circuit Stimulation"

*Supplementary Table 1. Patient Variables.*

| Score/Variable | STAGG-2 | STAGG-3 | STAGG-4 | STAGG-5 |
|---|---|---|---|---|
| Age (5-year range) | 68-73 | 74-79 | 50-55 | 60-65 |
| Sex and Gender | Male | Male | Male | Male |
| Disease Duration | 2 years | 3 years | 7 years | 4 years |
| Disease Subtype | MSA-P | MSA-P | MSA-P | MSA-P |
| UMSARS Hx | 20 | 18 | 23 | 27 |
| UMSARS Ex | 12 | 17 | 27 | 34 |
| COMPASS-31 | 57 | 31 | 30 | 33 |
| RBD Dx | No formal Dx | No formal Dx | No formal Dx | No formal Dx |
| RBDSQ | N/A | N/A | N/A | N/A |
| PDSS | N/A | N/A | N/A | N/A |
| ESS | N/A | N/A | N/A | N/A |
| Stimulants (current or prior use of amantadine or modafinil) | No | No | Not current (prior trial of amantadine, discontinued) | No |
| GFQ | 21 | 12 | 27 | 28 |
| FOGQ | 12 | 10 | 17 | 17 |
| PDQ-8 | N/A | N/A | N/A | N/A |
| ADL | 80% | 70% | 50% | 50% |
| EQ-5D VAS | 50 | 70 | 30 | 37 |
| Zarit Burden | | | | |
| ACEI-III | - | 100 | 93 | 93 |
| MOCA | 25 | - | - | - |
| HAD Scores | HADS-A: 10<br>HADS-D: 7 | HADS-A: 8<br>HADS-D: 4 | HADS-A: 5<br>HADS-D: 13 | HADS-A: 2<br>HADS-D: 9 |
| L-Dopa post-surgically | No | Yes | Yes | Yes |
| L Dopa cumulative dose (if yes) | - | 300mg DA | 800mg DA | 100mg DA |

---

*Abbreviations: MSA-P: Multiple system atrophy- parkinsonian type, UMSARS: Unified Multiple System Atrophy Rating Scale, COMPASS-31: Composite Autonomic Symptom Score-31, RBD: REM-Sleep Behaviour disorder, RBDSQ: REM Sleep Behavior Disorder Screening Questionnaire, PDSS: Parkinson's Disease Sleep Scale, ESS: Epworth Sleepiness Scale, GFQ: Gait and Falls Questionnaire, FOGQ: Freezing of Gait Questionnaire, PDQ-8: Parkinson's Disease Questionnaire, ADL: Activities of Daily Life, EQ-5D VAS: EuroQuol-5D Visual Analogue Scale, ACE I-III: Addenbrooke's Cognitive Examination I-III, MOCA: Montreal Cognitive Assessment, HADS, DA: Dopamine, Dx: Diagnosis, N/A: Not Available.*

### Supplementary Methods

*Baseline Data acquisition and Pre-processing*

All data were recorded using a TMSi Refa system (TMS International, Oldenzaal, Netherlands) at a sampling rate of 2048 Hz. An electrode cap (Easycap, Herrsching, Germany) was fitted to the head size of each participant, designed for EEG data to be recorded from sixty-four silver/silver chloride electrodes placed according to the extended 10/20 system. Furthermore, a ground electrode was positioned on the midline centrally while the common channel average was used as acquisition reference.

Due to the presence of entry wounds at the implantation sites, we modified electrode placement to leave these sites undisturbed. Additionally, we decided to avoid mastoid electrodes altogether since their placement interfered with comfort due to the externalised DBS electrodes in one patient. Therefore, for the modified recording electrode setup, a Cz re-referencing electrode was selected for sleep scoring in all cases to ensure homogeneity across recordings. Electro-occulogram (EOG) was recorded from bipolar electrodes placed above and below the outer canthi of right and left eye respectively, while electrocardiogram (ECG, to assess/monitor for any cardiac events) and chin electromyogram (EMG) were also placed as per AASM guidelines [1] .

All analyses were carried out in MATLAB using bespoke in-house scripts, incorporating the FieldTrip [2] toolbox. Individual channel data were de-trended to remove drift and filtered (high-pass Butterworth filter (3rd order) set at 0.2Hz and low-pass at 180Hz, with band-stop for 50Hz line noise and its harmonics) prior to any further analyses.

*Sleep Fragmentation and Index Calculation*

Sleep fragmentation was assessed per patient, after pre-operative baseline night and post-operative recovery night were visualised and scored as per AASM criteria [1] . Both nights were inspected (where pre-operative baseline was available) to ensure whether sleep depth was affected by implantation/stun effects. Fragmentation was first examined based on number of spontaneous wake-scored epochs occurring after the first sleep episode, defined as 'Wake after Sleep Onset' and scored based on the Pittsburgh Sleep Quality Index Scoring Algorithm [3]. Based on the Pittsburgh Sleep Quality Index scoring algorithm, WASO ≤ 15% suggested minimally disrupted sleep while > 25% indicated highly disrupted sleep. WASO was therefore categorized into three groups ≤ 15%, 15-25%, and > 25% to characterise sleep disturbance in this cohort.

In order to assess sleep continuity further, in addition to epochs scored as 'wake' based on AASM criteria, we also investigated the presence of ISARs as sleep disrupting events and subsequently calculated ccontinuous (>10mins) sleep episodes occurring after the first epoch of non-REM sleep (NREMS). After continuous sleep periods and frank epochs of wake after sleep onset (WASO) were identified, relevant scores were calculated and used in later analyses and evaluations. These consisted of three indices. Firstly a WASO (%) index, calculated as the percentage (%) of wake epochs occurring after the first sleep episode, divided by the total number of epochs from this episode until the first wake epoch with which sleep was ended for the night. Then a continuity (%) index, calculated as the percentage (%) of epochs in continuous sleep episodes (as defined previously, exceeding 10 minutes in duration) divided by the total number of epochs from the first sleep episode.

*Stimulation Trial Data Pre-processing*

Trial data (EEG and LFP) were visually inspected post-hoc to identify artefacts (motion, stimulation-related etc) and optimal frontal channels for analyses, as well as to formally score sleep depth of the epochs preceding stimulation. Exclusion criteria of trial data from further analyses included delivery outside of slow-wave sleep, excessive artefact presence during the post-stimulation period and/or any trials where stimulation duration exceeded standardised protocol parameters. Trial data were assessed with regards to presence of wake, ISARs and stage shifts. We were able to record EEG and contralateral LFPs during unilateral PPN stimulation, while we had no capacity to stream LFPs from the device while stimulating bilaterally.

In more detail, for each individual stimulation trial, channel data were visualised after de-trending and filtering (as described above). Frontal and prefrontal channels were assessed for signal quality and presence of artefact within the three AASM sleep epochs preceding and following the stimulation period, identified both by experimental records and the presence of stimulation artefact. EEG channels with minimal artefact were then selected and re-referenced (central reference of Cz, FC1 or FC2). In addition, PPN LFP channels where maximal gamma modulation had been identified during passive recordings were assessed for signal quality and re-referenced in the same manner as in prior baseline analyses.

*Brainstem Power Modulation by putative UP- and DOWN-State*

From the EEG channels showing maximal SWA power and during these scored NREM2 and NREM3 epochs, slow waves were detected with the following criteria in addition to frequency (0.5-4Hz): peak-to-peak amplitude exceeding 50 μV as well as train (SWA trains or ‘snippets’) duration of 5sec. Such detected ‘snippets’ were visually inspected to ensure signal quality prior to phase calculations. In addition, we selected control state ‘snippets’ from epochs scored as quiet wakefulness (eyes closed), where slow-wave activity was not detected. These control ‘snippets’ were also visually inspected for signal quality.

We considered one complete UP to DOWN-state cycle (reflecting changes between excitation and inhibition) to be 2π, first finding all zero-crossing points from the bandpass-filtered EEG data. The points corresponding to the negative peak of the inferred 'DOWN-State' were assigned phase -π while positive maxima (peak 'UP-state', with the constraints mentioned in the body of the main article) were assigned phase π. The phases of all other time points in the oscillatory cycle were determined through linear interpolation between –π and 0 (DOWN-State to zero-crossing) or between 0 and π (zero-crossing to UP-state). We further divided each cycle into 18 non-overlapping bins spanning from –π to π.

To calculate a 'phase/power modulogram' we then decomposed filtered bipolar LFP channels in the time–frequency domain by applying continuous complex Morlet wavelet transforms, with a linear frequency scale ranging from 1 to 95 Hz and a linearly spaced number (4–8) of cycles across all calculated frequencies. The LFP power spectral densities (PSDs) were further estimated by averaging the wavelet power across the time periods of interest and then normalized by the sum of the whole frequency band between 1 and 95 Hz. To quantify a modulation index (MI) per frequency, the mean power within each cycle phase bin per frequency was then normalized against the sum of all bins to give a percentage of power for each phase bin. Then, the MI was defined based on the Kullback–Leibler (KL) distance between the phase-locked power at each frequency and a uniform distribution as follows [4,5] :

$$\mathrm{MI} = \frac{D_{\mathrm{KL}}(P,U)}{\log(N)}$$

$$D_{\mathrm{KL}}(P,U) = \sum_{j=1}^{N} P(j) \log\left[\frac{P(j)}{U(j)}\right]$$

where *P* and *U* indicate the phase-locked power distribution at a specific frequency and a uniform distribution, respectively. N indicates the number of bins (i.e., 18). Here the KL distance is used to quantify differences between two distributions, which has the property that $D_{KL}(P,U) \geq 0$, and $D_{KL}(P,U)=0$ if and only if $P = U$ (i.e., when the distributions are the same). Large deviations from the uniform distribution result in a large modulation index.

Additionally, the mean power within each phase bin was calculated for each frequency and normalized against the mean of all bins, in order to give a percentage of power for each phase bin as an additional estimate of power changes across the SWA cycle.

The control wake state was not characterized by phasic changes in UP/DOWN State activity and had negligible SWA power content, therefore we calculated a 'sham' phase based on a random 'transition' point in the band-passed signal. The LFPs were then phase-locked to this random 'sham' phase for calculation of power modulation indices.

*Functional Connectivity Analyses*

For the two channels i and j let there be time series $x_i(t)$ and $x_j(t)$, with complex Fourier transforms $\hat{x}_i(f)$ and $\hat{x}_j(f)$. Therefore, the cross-spectrum of channels i and j is calculated as

$$S_{ij}(f) \equiv \langle \hat{x}_i(f)\hat{x}_j^*(f) \rangle$$

with $\hat{x}_i(f)$ the Fourier transform of the time series $x_i(t)$ recorded at channel i, $*$ denoting complex conjugation and $\langle\ \rangle$ expectation value. Complex coherency is defined based on the above cross-spectrum as

$$C_{ij}(f) \equiv S_{ij}(f)/(S_{ii}(f)S_{jj}(f))^{0.5}$$

The imaginary coherence can be obtained by projecting the complex-valued coherency onto the imaginary axis (additionally see [6] ). The imaginary coherence estimates were therefore calculated individually for each combination between re-referenced lead contacts and/or EEG electrodes of two areas of interest, in a coherence matrix specific to condition (quiet wakefulness, slow-wave sleep). The magnitude of the imaginary component was used to indicate the degree of co-variability between the two signals over different frequency ranges in a manner least affected by volume conduction and artefactual changes. We used a multi-taper method based on the FFT calculated during the previous step, for the FOIs denoted before.

We also sought to establish the effective connectivity between subcortical and cortical areas of interest in order to assess state-dependent changes in directionality of information flow. To this end we used Granger causality [7] , based on a multivariate autoregression model.

Formally, Granger causality determines whether the predictions given by the autoregression of y:

$$y_t = a_0 + a_1 y_{t-1} + \cdots + a_n y_{t-n} + \varepsilon_t$$

are improved by inclusion of lagged values of x:

$$y_t = a_0 + a_1 y_{t-1} + \cdots + a_n y_{t-n} + b_p x_{t-p} + \cdots + b_q x_{t-q} + \varepsilon_t$$

Since a parametric estimate of Granger causality is often difficult to calculate due to over-fitting the autoregression model, we also calculated a non-parametric measure using Fieldtrip function ft_connectivityanalysis() [2]. We compared different combinations of time window (twin) and model order (MO) over multiple iterations, to determine the best data fit, tailored to frequency of interest and both with and without down-sampling at 200Hz. The optimal parameters were selected based on which one provided the maximal values of the parametric and non-parametric model order, since they are inversely related to prediction error.

*Phase-Locking Value Calculation*

We first considered re-referenced EEG signals to minimize stimulation artefacts (*Supplementary Figure 1*). For each individual trial, data were visualized across individual channels to select both channel with maximal artefact (for trigger detection) as well as appropriate re-referencing pairs. The number of stimulation pulses was computed by obtaining individual peaks using the Matlab function ‘findpeaks’ on the raw acquired signal of the channel with maximal stimulation artefact as input signal vector for the detection. The result was verified both by visual examination of each trial as well as by ensuring that the number of peaks detected after threshold specification corresponded to the stimulation frequency that had been experimentally applied (*Supplementary Figure 2)*. Re-referenced signals were then bandpass filtered, centered around the half-width of the band of interest. Other comparative bands of interest in addition to the aforementioned gamma ranges included alpha and beta oscillations (at  [8 12) and [12 30] Hz respectively). In addition, control signals of pink noise were generated, for the same duration per each trial.

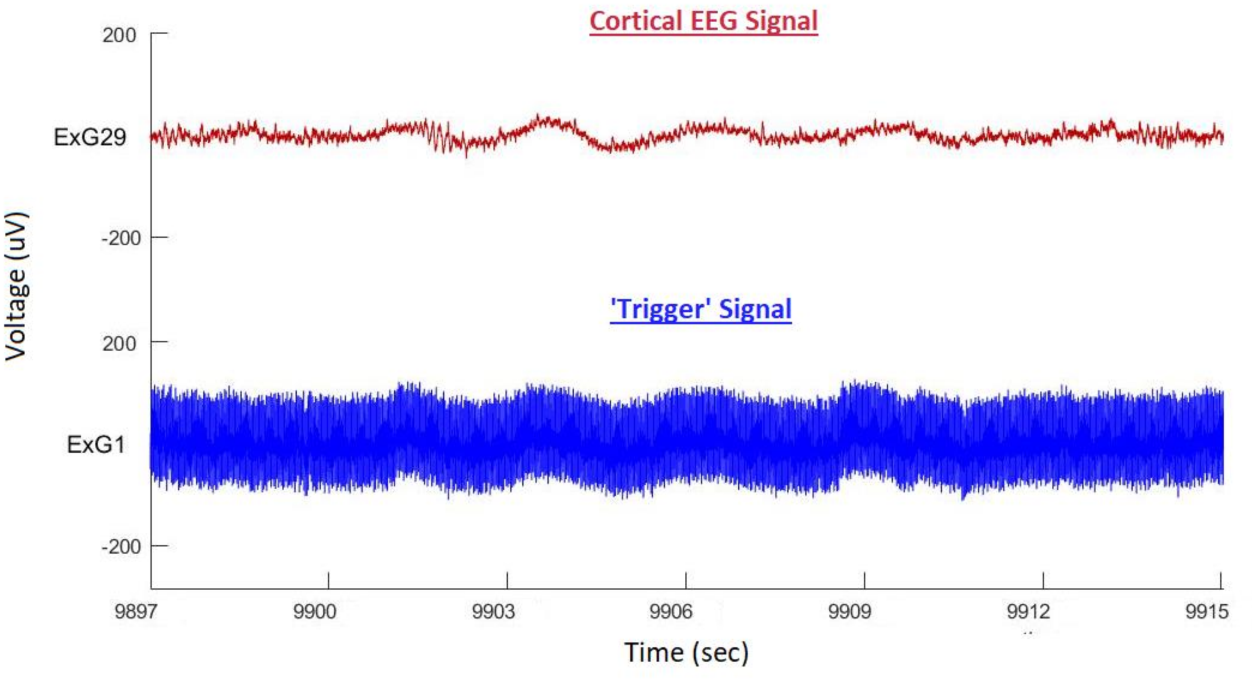

*Supplementary Figure 1. Artefact rejection. Re-referenced pair (channel 29 –channel of interest- in a dipole with channel 12), compared with 'trigger' signal, later used for peak detection (channel 1, un-rereferenced). Note how stimulation artefact is not present in the dipole containing the channel of interest, where oscillatory activity can also be visually discerned.*

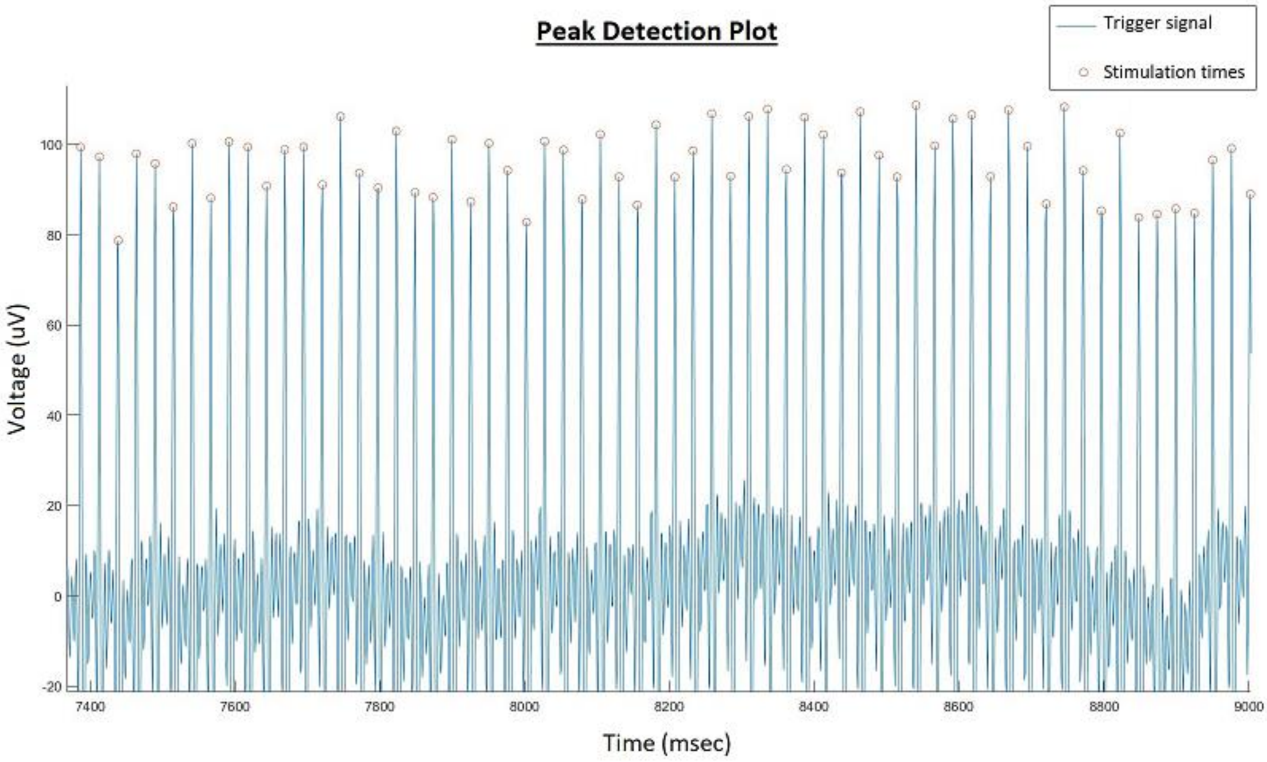


*Supplementary Figure 2. Stimulation artefact's peak detection method. We wished to verify that, for a minimum prominence (threshold) set, stimulation times were correctly identified based on the artefact present in the 'trigger' channel signal.*

In each case of both trial and control (pink noise) data, the Hilbert phase $\psi(t)$ was obtained from the bandpass filtered signal.

To quantify synchronisation to the stimulation frequency $f_{stim}$, we calculated for each trial and channel

$$\text{PLV 1:1} = \frac{1}{N}\left|\sum_{k=1}^{N} e^{i\psi(t_k)}\right|,$$

where $N$ is the number of stimulation pulses in one trial, $t_k$ is the time of the k$^{th}$ stimulation pulse within the trial, and $|.|$ denotes the complex modulus. PLV 1:1 quantifies the phase concentration at the time of stimulation, considering all stimulation pulses. To quantify synchronisation to subharmonics of the stimulation frequency $f_{stim}/p$, we calculated for $p = 2$ and $p = 4$, for each trial, channel, and condition

$$\text{PLV } 1{:}p = \frac{1}{\left\lfloor \frac{N}{p} \right\rfloor} \left| \sum_{k=1}^{\lfloor N/p \rfloor} e^{i\psi(t_{pk})} \right|,$$

with $\lfloor . \rfloor$ the floor function. PLV 1:$p$ only considers every $p$ stimulation pulses ($t_{pk}$ is the pk$^{th}$ stimulation pulse in the trial), and therefore characterises phase concentration at the subharmonic 1:$p$ of the stimulation frequency. For each channel and condition, PLV 1:1 and PLV 1:$p$ were averaged across trials (*Supplementary Table 2 A, B)*.

*Supplementary Table 2. Mean PLV values per stimulation protocol (including sub-harmonics).*

A. 40Hz Stimulation Frequency: mean PLV values

| PLV pair with endogenous oscillation: | Frontal | Occipital |
|---|---|---|
| Alpha with stimulation frequency (±SD) | 0.0483 (±0.1153) | 0.0191 (±0.0492) |
| Beta with stimulation frequency (±SD) | 0.0878 (±0.0949) | 0.0390 (±0.0527) |
| Gamma with stimulation frequency (±SD) | 0.6061 (±0.2917) | 0.4914 (±0.1659) |
| Beta with half-harmonic of stimulation frequency (±SD) | 0.0942 (±0.0923) | 0.0448 (±0.0629) |
| Alpha with quarter-harmonic of stimulation frequency (±SD) | 0.0774 (±0.1095) | 0.0421 (±0.0436) |

B. 100Hz Stimulation Frequency: mean PLV values

| PLV value with endogenous oscillation: | Frontal | Occipital |
|---|---|---|
| Alpha with stimulation frequency (±SD) | 0.0204 (±0.0326) | 0.0031 (±0.0067) |
| Beta with stimulation frequency (±SD) | 0.0352 (±0.0530) | 0.0077 (±0.0096) |
| Gamma with stimulation frequency (±SD) | 0.0364 (±0.0186) | 0.0300 (±0.0164) |
| Beta with quarter-harmonic of stimulation frequency (±SD) | 0.0448 (±0.0608) | 0.0143 (±0.0098) |

*Supplementary Statistics*

Firstly, we used the Kolmogorov-Smyrnov method to test for normality of residuals of the power-in-band for the analysis model. Since the assumption was violated, a Box-Cox transformation was used on the data prior to ANOVA analyses examining differences between protocols in the frequency bands of interest. Again we used a Bonferroni correction for all multiple comparisons. We also deployed the Wilcoxon Rank Sum test to compare differences in PLV between simulated pink noise trials and stimulation trials, across different cortical regions and oscillatory frequencies, within the same stimulation frequency group (40Hz and 100Hz) and their sub-harmonics. In more detail, a two-way ANOVA (parameters: frequency of stimulation and frequency of endogenous oscillations) was used per region (frontal, occipital), on the data post transformation and with Bonferroni correction. An additional three-way ANOVA analysis was used for the comparison between pooled PLV values (for both frontal and occipital EEG channels) as well as individual cortical regions, across the different stimulation frequency groups and sub-harmonics thereof (post transformation, with Bonferroni correction).

With regards to the regional, two-way ANOVA analyses, frontally there was no statistically significant difference between PLVs of both protocols with regards to endogenous alpha oscillations ($p= 0.5286$, CI= -0.3814 to 2.2692). Occipitally however, phase-locking was significantly higher between the 40Hz stimulation protocol and endogenous cortical alpha activity ($p< 0.0001$, CI= 1.2283 to 3.8630). With regards to beta oscillations, the 40Hz protocol outperformed the 100Hz protocol with regards to higher phase-locking with cortical activity in both regions (frontally $p= 0.0099$, CI= 0.1719 to 2.2439 and occipitally $p= 0.0031$, CI= 0.2869 to 2.3424). The degree of phase-locking of the 40Hz protocol was also significantly higher than that of the 100Hz protocol with regards to gamma oscillations ($p<0.0001$ in both regions, all p values with Bonferroni correction). In the three-way ANOVA examining the pooled PLV values across both regions, the 40Hz protocol (and its subharmonics) showed significantly higher PLV values with regards to both alpha, beta and gamma cortical oscillations ($p<0.0001$ with Bonferroni correction) (see *Supplementary Table 3* for detailed report of all ANOVA statistical parameters).

*Supplementary Table 3. PLV ANOVA statistics.*

A. Regional two-way ANOVA: Frontal

| Source | Sum Sq. | d.f. | Mean Sq. | F | Prob>F |
|---|---|---|---|---|---|
| stim_vect | 81.409 | 1 | 81.4093 | 44.22 | <0.0001 |
| freq_vect | 111.844 | 2 | 55.9221 | 30.37 | <0.0001 |
| stim_vect*freq_vect | 17.48 | 2 | 8.74 | 4.75 | 0.01 |
| Error | 270.636 | 147 | 1.8411 | | |
| Total | 511.492 | 152 | | | |

*Abbreviations: stim_vect: stimulation protocols (40Hz, 100Hz). freq_vect: endogenous oscillation frequency (alpha, beta, gamma). d.f: degrees of freedom.*

B. Regional two-way ANOVA: Occipital

| Source | Sum Sq. | d.f. | Mean Sq. | F | Prob>F |
|---|---|---|---|---|---|
| stim_vect | 135.02 | 1 | 135.02 | 89.06 | <0.0001 |
| freq_vect | 199.678 | 2 | 99.839 | 65.86 | <0.0001 |
| stim_vect*freq_vect | 16.203 | 2 | 8.102 | 5.34 | 0.0059 |
| Error | 187.98 | 124 | 1.516 | | |
| Total | 537.256 | 129 | | | |

*Abbreviations: stim_vect: stimulation protocols (40Hz, 100Hz). freq_vect: endogenous oscillation frequency (alpha, beta, gamma). d.f: degrees of freedom.*

C. Summary three-way ANOVA

| Source | Sum Sq. | d.f. | Mean Sq. | F | Prob>F |
|---|---|---|---|---|---|
| stim_vect | 210.38 | 1 | 210.381 | 120.21 | <0.0001 |
| freq_vect | 301 | 2 | 150.502 | 86 | <0.0001 |
| location_vect | 17.92 | 1 | 17.923 | 10.24 | 0.0015 |
| stim_vect*freq_vect | 27.69 | 2 | 13.846 | 7.91 | 0.0005 |
| stim_vect*location_vect | 2.3 | 1 | 2.3 | 1.31 | 0.2526 |
| freq_vect*location_vect | 3.85 | 2 | 1.924 | 1.1 | 0.3345 |
| Error | 477.77 | 273 | 1.75 | | |
| Total | 1075.94 | 282 | | | |

*Abbreviations: stim_vect: stimulation protocols (40Hz, 100Hz). freq_vect: endogenous oscillation frequency (alpha, beta, gamma).* location_vect*: main cortical region (frontal, occipital). d.f: degrees of freedom.*

To accurately identify frequency bands with significant state-related functional connectivity changes, we used a non-parametric cluster-based permutation procedure while controlling for multiple comparisons [8] . To compare the group averages between conditions (quiet wakefulness and SWS, as well as pre-stimulation and post-stimulation or their 'SHAM' analogues), the condition labels of the original samples were randomly permutated 1000 times resulting in a null hypothesis distribution of 1000 samples. The permutation distribution mean and SD were then used to z-score the original un-permuted data, obtaining a p value for each data point. Suprathreshold clusters were obtained for the original unpermuted data and each permutation sample by setting a pre-cluster threshold ($p < 0.05$). Statistical significance was considered if the absolute sum of the z-scores within the original suprathreshold clusters exceeded the 95th percentile of the 1000 largest absolute sums of z-scores from the permutation distribution (i.e., at $p < 0.05$).

In order to assess the significance of power modulation indices for LFP contact pairs linked to UP- and DOWN-State, we used the same non-parametric method of obtaining suprathreshold clusters for unpermuted data. The null hypothesis comparator distribution here was obtained by randomly shuffling the phase for each individual slow-wave cycle. The phase vector was re-calculated by selecting a random transition point for division of phase (at any point during UP- or DOWN-State transitions), then randomly shuffling phase for calculation of a new modulation index. This was repeated 1000 times to obtain a permutation distribution with 1000 samples as in the prior description. For comparisons of modulation indices in the control wake state, the additional null hypothesis vector was calculated by a second random point for phase division in a similar way.

To explore whether bottom-up (from PPN to frontal cortex) information flow during quiet wakefulness and sleep may be linked to sleep fragmentation, a factorial ANOVA was computed between directional Granger causality (GC) values, sleep continuity and WASO indexes for each macro-state (wake and sleep). We also compared for general inter-patient differences in GC values (regardless of cause) with an ANOVA with sleep, wake and patient anonymised ID as factors. A Bonferroni correction was used in all analyses to counter for multiple comparisons.

### Supplementary Results

#### ***Gamma and Beta Brainstem Oscillations are modulated by Cortical UP- and DOWN-State transitions***

We further sought to quantify changes correlated to UP/DOWN cycles of theta, alpha, beta and gamma frequency power-in-band, additionally separating gamma into low (below 45Hz) and high (above 55Hz) gamma bands. Specifically, for alpha and beta power, we selected the bipolar LFP channels with the maximum power during DOWN state while for theta, low and high gamma power, we selected the bipolar LFP channels with the minimum power during DOWN state. We subsequently quantified the average power-in-band from the selected bipolar LFP channels and compared them between the two parts of the slow wave cycle. Beta power was significantly higher during the DOWN-State (p=0.04788, t=2.3983), theta power was higher during the peak of the UP-state (p= 0.0018, t=-4.8744) while high gamma power significantly increased during transition to UP-State (p=0.002666, t= -4.5406). The differences in alpha and low gamma bands did not reach statistical significance.

#### *Bottom-Up Beta and Gamma Information Flow during Sleep correlates with Sleep Fragmentation*

When differences in gamma and beta GC for both sleep and wake were examined between patients, we noted that there was no significant difference in wake GC values between any of our patients, across all examined frequency ranges (p>0.1 in all cases). However, we noted that in all cases bottom-up information flow increased significantly across the beta and gamma ranges during SWS (p<0.00001 in all cases, after Bonferroni correction), suggesting the presence of a common process active in all patients. We also observed that overall, the highest values of beta and gamma GC during SWS were present in the case with the poorest capacity for sleep maintenance as signified by lowest sleep continuity score and highest percentage of wake after sleep onset.

To further explore circuit connectivity during the two states, as well as identify any links between PPN activity and fragmented sleep, we additionally looked at changes in information flow directed from brainstem to cortex. We therefore calculated Granger Causality (GC) during SWS and quiet wake epochs, analysing it as described previously (*Methods*). We subsequently assessed sleep fragmentation and continuity in our cohort. We noted that in all cases there was poor sleep continuity; maximally only 1/3 of the sleep period (from first to last sleep epoch) consisted of continuous sleep epochs uninterrupted by ISARs, wake and significant artefact (*Supplementary Table 2*).

*<u>Supplementary Table 4</u>. <u>Sleep fragmentation indexes per patient.</u> Calculated for the post-operative night (after recovery from Stage 1) when brainstem LFPs were sampled for analysis with simultaneous polysomnography. WASO: Wake After Sleep Onset.*

| **Patient ID** | **WASO Index (%) ł** | **Continuity index (%) *** |
|---|---|---|
| **STAGG-2** | **24.48** | 16.72 |
| **STAGG-3** | **13.75** | 25.21 |
| **STAGG-4** | **12.44** | 27.80 |
| **STAGG-5** | **34.65** | 16.16 |
| ł Percentage % of wake epochs/total Nº epochs from the first NREM episode<br>*Percentage % of epochs in continuous sleep episodes (exceeding 10 mins in duration)/ total Nº epochs from the first NREM episode. | | |

We then calculated WASO indices per patient and subsequently pooled all wake and sleep epochs according to the degree of sleep disruption identified. In two of our patients, WASO was < 15% (suggesting minimal presence of frank arousals after sleep onset), while one patient had moderate sleep disturbance (defined as WASO 15-25%) and one patient's WASO exceeded > 25% signifying severe fragmentation. When epochs were pooled per category, we noted that there were no statistically significant differences between degree of sleep disturbance (WASO category) and values of wake bottom-up information flow (p>0.1 in all cases). However, we saw that increasingly pathological WASO was correlated to higher values of beta GC during SWS, with each tier/group having sequentially higher GC values than the prior (less severe) one (p<0.0001 with Bonferroni correction for all case comparisons). With regards to gamma GC during SWS, all groups had higher values compared to those during wakefulness (p<0.0001 with Bonferroni correction for all case comparisons). However, here the relationship between WASO index severity and increase in GC during SWS was less clear; although the lowest WASO group had the lowest relative values of GC during sleep, the increase was not sequential for the next two groups.

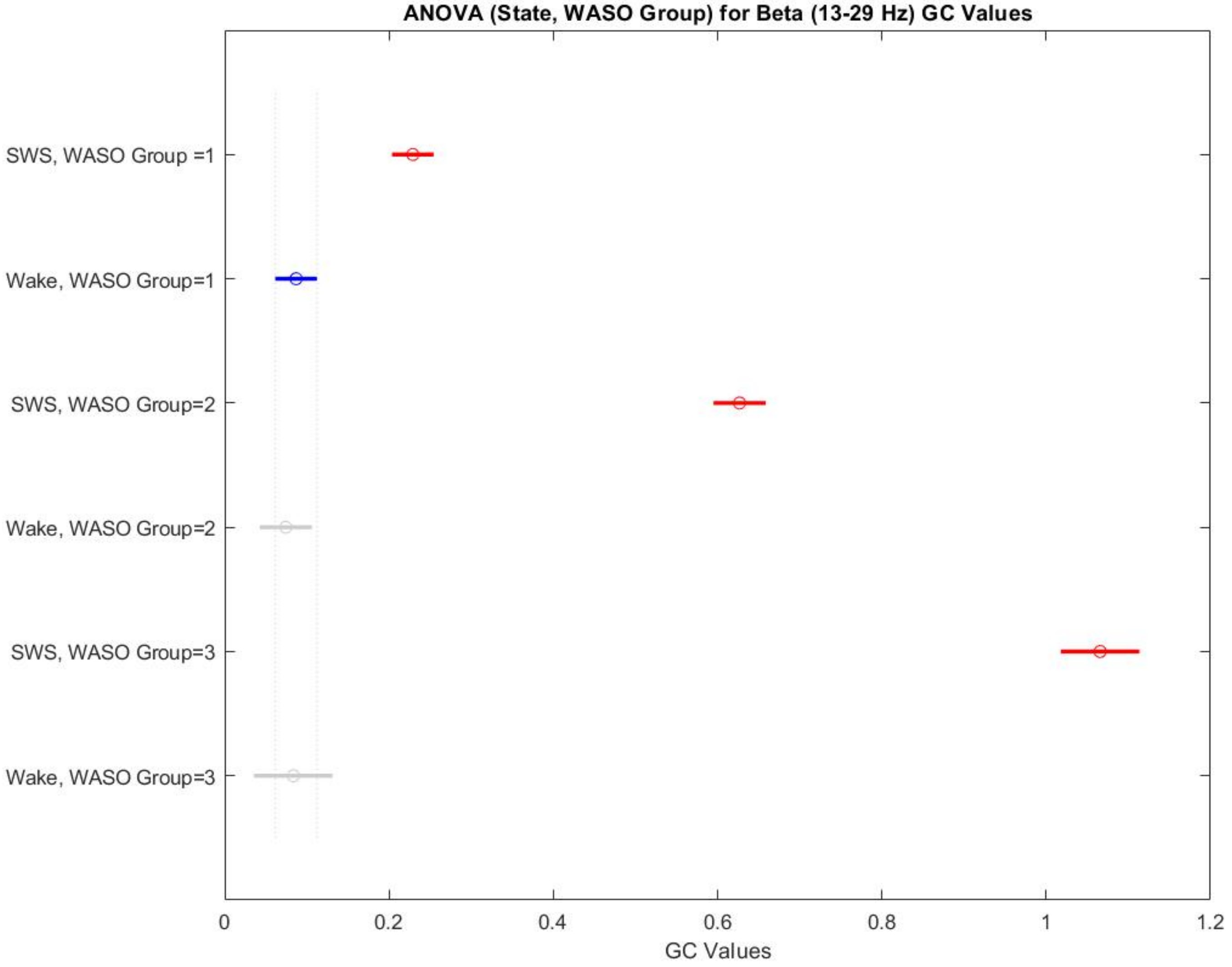


*Supplementary Figure 3. Bottom-up information flow from brainstem to cortex increases during SWS in the beta range, correlating with degree of sleep disruption (WASO index). WASO Group 1 characterised by <15% of wake after sleep onset epochs, WASO Group 2 15%-15% indicating moderate sleep disruption and WASO Group 3 wake epochs >25% indicating severe sleep disruption. Groups between dashed lines: no statistically significant difference between sub-groups (here shown in blue and grey). Red: statistically significant difference between sub-groups. Statistical values for all comparisons reported in Supplementary Table 1.*

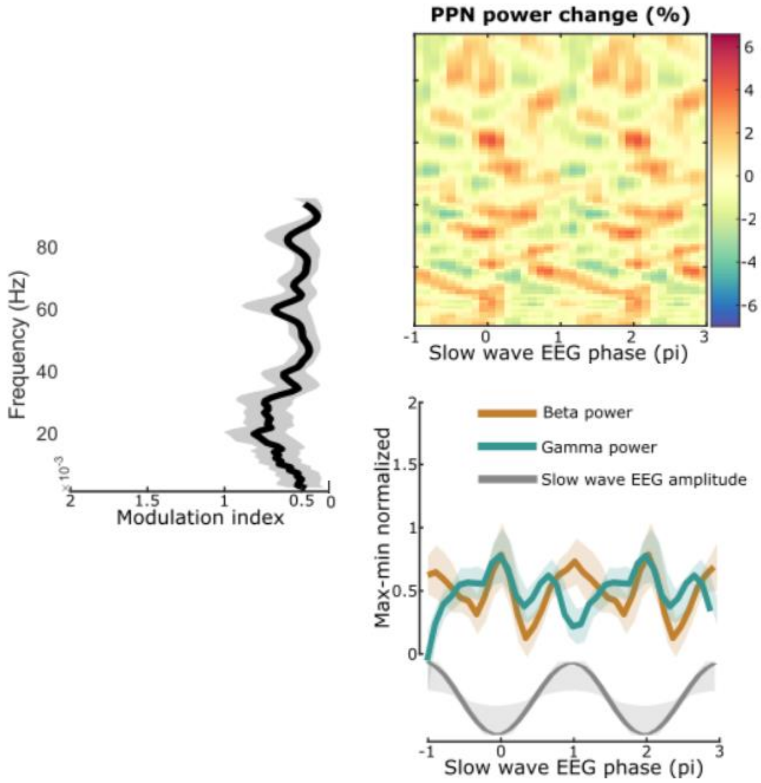


<u>*Supplementary Figure 4.*</u> <u>*During wakefulness, beta and gamma oscillations show no modulation by the equivalent of the SWA cycle.*</u> *Compare with Figures 3 and 4 in SWS.*

#### *Stimulation Trial Delivery*

The shorter sleep duration and lighter sleep depth as a consequence of intermittent brainstem stimulation during the 'STIM' nights resulted in limited capabilities in multiple trial collection per night. Additionally, since we had pre-specified that trials would only be included in analyses if the preceding cortical state was comparable in terms of sleep depth and slow-wave activity, this limited the number of trials we could collect per patient. Where length of stay and patient tolerance rendered it feasible, this constraint was mitigated by trial collection over two subsequent nights.

One of the patients experienced an episode of confusional arousal during their first stimulation night (with agitation and associated brief hallucinations upon abrupt post-stimulation awakening). Therefore, the recording of further trials was discontinued for the night. This was a new behaviour as per their report; subsequently no further overnight experimental high frequency (100Hz) trials were delivered during their participation, while furthermore their clinical settings were tested and adjusted so that stimulation remained OFF at night. Another limitation with regards to bilateral trial data collection came from the fact that one other case contracted a hospital-acquired pneumonia during their stay, with subsequent deterioration and transfer to the Intensive Care Unit. No further experimental trials were therefore feasible (save for initial unilateral trial data collection, on a night with severe sleep fragmentation), while this case was also discharged to a local hospital before the second stage of their operation with significant frailty. Therefore, there was no feasibility of performing bilateral stimulation trials for this study.

Trials were therefore unequally distributed across cases, due to the aforementioned constraints. We initially sought to assess stimulation effects on a number of comparably distributed cases across our four implanted patients, with the additional clause of stage/slow wave depth equivalence within- and across-subjects. One could thus argue that should we have pooled all trials that fulfilled post-delivery inclusion criteria, slight differences in implantation depth or patient characteristics might skew the result. Overall, we applied DBS on 45 cases of discrete trials (total of unilateral and bilateral trials of both 40Hz and 100Hz) during live scoring. For further analyses, twenty matched stimulation trials of comparative laterality and appropriate duration, evenly distributed across cases and delivered during epochs scored as SWS post-acquisition, were therefore selected (ten at 40Hz and ten at 100Hz), with eighteen SHAM trials all delivered during SWS for comparison.

However, when the preceding three AASM epochs were examined, sleep depth had fluctuated over this time period in five cases (f.ex. sequentially NREM2, NREM1, W, NREM1, NREM2). When slow wave power was examined, these trials with prior evidence of stage instability (fragmentation/fluctuating depth) differed significantly from other pre-SHAM/STIM cases and were therefore excluded. The residual trials (ten trials at 40Hz, sixteen SHAM trials, seven trials delivered at 100Hz) were then again compared in terms of pre-stimulation cortical state in terms of slow wave activity (SWA) power and there was no statistical significance between the groups (*Figure 6A*). We ensured that the final number of SHAM trials as well as stimulation trials for each frequency was matched within and between cases (for instance four cases of SHAM trials, four cases at 100Hz and three at 40Hz from the same patient, all with comparable sleep depth).

Lower cortical gamma (30-45Hz) increased post-stimulation in comparable trials, with $p<0.0001$ for both 40 Hz (CI -0.0207 to -0.0101) and 100Hz (CI -0.0187 to -0.0067) while there was no significant increase in low gamma after the SHAM trials ($p = 0.3428$, CI -0.0009 to 0.0072). This was also true for higher cortical gamma (55-70Hz), with $p<0.0001$ for both 40 Hz (CI -0.0228 to -0.0117) and 100Hz (CI -0.0218 to -0.00925) and no significant increase in low gamma after the SHAM trials ($p = 1$, CI -0.0042 to 0.0043). The ‘physiological’ stimulation (40Hz) also resulted in a very significant increase in alpha power during the post-stimulation period ($p< 0.0001$, CI -0.0995 to -0.05389). A significant increase in alpha was also noted after the ‘supra-physiological’ gamma protocol was delivered ($p = 0.0047$, CI -0.0598 to -0.0061). In the case of the SHAM trials, there was no change in alpha power occurring after the ‘stimulation’ was applied ($p = 1$, CI -0.0256 to 0.0108).

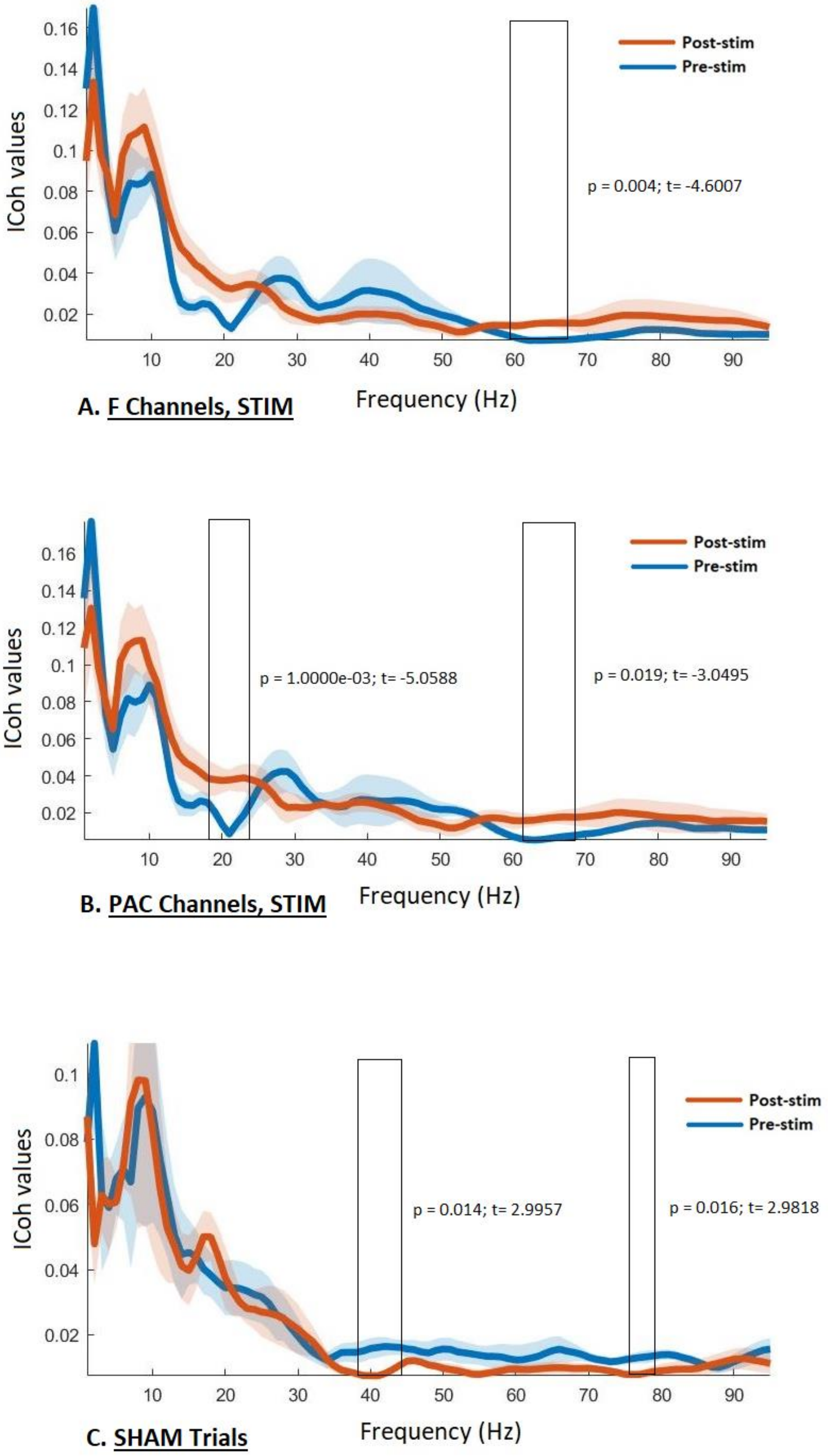


*<u>Supplementary Figure 5</u>. <u>Changes in Imaginary Coherence (ICoh) between PPNR LFPs and EEG, after stimulation (STIM) and SHAM trials</u>. A. For all frontal and prefrontal EEG channels, the post-stimulation period is characterized by a significant increase in high gamma coherence (frequency encased by rectangular shape). B. ICoh between PPNR LFPs and*

*prefrontal channels showing maximal slow wave power (used in phase-amplitude coupling (PAC) during the previous chapter, hence denoted as PAC in the graph). An additional finding here is increase in ICoh in the beta frequency range (close to the stimulation half-harmonic). C. ICoh between all channels (frontal/prefrontal) and PPNR LFPs. Here, we note an opposite phenomenon –decreases in ICoh between brainstem and cortex, in the gamma range, as sleep stage in the majority of cases progressed in depth or stabilized. This change occurs in the low gamma (39-44Hz) and high gamma (75-79 Hz). Please refer to the text and figures for p and t values.*

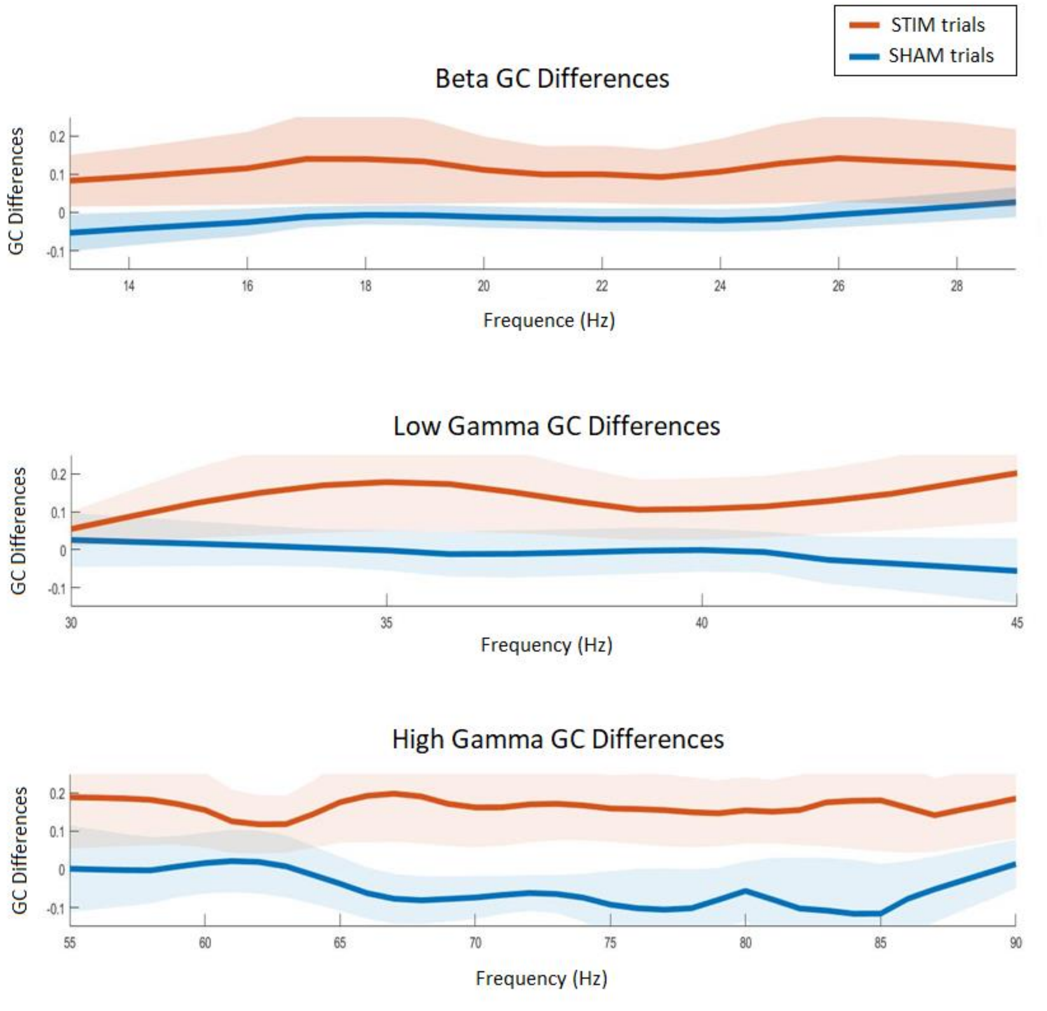


*<u>Supplementary Figure 6</u>. <u>Comparisons between SHAM and stimulation trial changes in information flow (GC)</u>. Negative values denote decreases in information flow (present in the SHAM cohort post 'stimulation' when sleep depth does not decrease). Positive values denote increases in bottom-up information flow (and are present in the STIM –stimulation- cohort). Levels of statistical significance and associated metrics are described in the text.*

We focused on the stimulation periods of both 40Hz and 100Hz trials, as selected above. We also sought to identify any potential regional differences in this relationship, given that SWA power and generation differs and is mainly frontal, as discussed earlier in this thesis.

We examined all EEG channels to identify stimulation artefacts, identifying the exact time of pulse delivery comparatively to endogenous cortical phase of the oscillations of interest. During this process, especially for the higher stimulation frequency where amplitudes were low (1mA and below) due to threshold maintenance, we had to discard further trials where the peaks of pulse delivery could not be reliably identified –despite the fact that a stimulation artefact was present in the power spectra, verifying the validity of the trial (Supplementary *Figure 7*). We did not try to remedy this by creating an artificial signal (stimulation frequency sinusoid) and timing it to the cortical EEG signal for two reasons. Firstly, since there is a built-in delay of milliseconds from triggering until stimulation delivery within the stimulation box or implanted stimulation generator (IPG) and secondly, since triggering was physician-operated. Although we might assume that an IPG built-in delay is relatively constant, there is a variability where any human element is introduced that could influence precision with regards to phase-relationships. This issue resulted in the exclusion of one of our patients, where stimulation artefact peaks were not reliably identified with neither visual nor automatic detection.

With regards to the cortical frequencies that we investigated, we focused on alpha (8-12Hz), beta (13-30Hz) as well as the part of the gamma range where the DBS frequency is included and where we saw significant decreases in connectivity with sleep stage progression in the SHAM trials (39-44Hz). The rationale being that the capacity of the DBS (imposed) oscillator to synchronise brainstem areas and –should PLV values support this- drive cortical areas in this frequency. We aimed to focus on lower frequency bands during the stimulation period, as opposed to very high gamma frequencies, mainly due to the fact that during DBS stimulation the EEG traces were often contaminated by high-frequency artefacts associated with motor activity (while we also excluded parts of the traces where significant artefacts would potentially compromise the accuracy of our analyses). We further explored coupling between the half-harmonic of the stimulation frequency and cortical beta (for a stimulation frequency of 40Hz), as well as its quarter-harmonic and cortical alpha and beta rhythms (stimulation frequencies of 40Hz and 100Hz respectively). All stimulation trials were compared to generated pink noise control trials, with which phase-locking values of the stimulation were also examined.

With regards to frontal and pre-frontal dipoles, we noted significantly higher phase locking values than noise between stimulation frequency and cortical oscillations in the beta ($p < 0.0001$, zval=4.9086) and gamma frequency ($p < 0.0001$, zval=5.5367). The half-harmonic of the stimulation also significantly phase-locked to cortical beta compared to the synthetic pink noise control signals ($p < 0.0001$, zval=4.0280). There was no statistically significant difference in phase-locking with endogenous beta between the stimulation frequency (40Hz) and its half-harmonic ($p = 0.5294$, zval=-0.6289). In this cortical region, we noted that the stimulation did not significantly couple to alpha, neither at the stimulation frequency (p = 0.4569, zval=0.7440) nor at the quarterharmonic (p = 0.8013, zval=-0.2517) compared to pink noise trials.

When occipital dipoles were examined separately, we saw again that there was significantly high phase locking with cortex in real (40Hz effects on cortical activity) stimulation trials than in control (40Hz effects on pink noise) trials, in the beta and gamma frequency range (p <0.0001, zval=4.8634 and p <0.0001, zval=5.1149 respectively). The half-harmonic of the stimulation frequency was also significantly phase-locked to cortical beta oscillations occipitally (p =0.0325, zval=2.1386).

Subsequently, we explored how pulses delivered during high ('supra-physiological') frequency stimulation trials (100Hz), during periods of comparable slow wave sleep depth, may be coupled to frontal and occipital cortical rhythms. As mentioned previously, due to the very low amplitudes delivered at this frequency to ensure that stimulation did not cause sensory/other disturbance, automatic peak identification of unilateral trials was extremely challenging. Therefore, for unilateral trials delivered at 100Hz, only data from two patients were of sufficient artefact strength to be analysed. This therefore left us with twelve frontal/pre-frontal and ten occipital dipoles of the same laterality (as well as the same laterality as the 40Hz trials), from trials delivered during sleep epochs with the same depth and SWA density as during the 40Hz trials, available for pooling in our respective analysis groups.

With regards to the frontal group, the most significant effect was with regards to phase-locking of the stimulation frequency to the cortical gamma oscillation band of interest, compared to pink noise signals (p <0.0001, zval=4.1434). Additionally, the stimulation frequency significantly locked to beta oscillations compared to noise (p =0.0003, zval=3.6451). Given that the half-harmonic of this stimulation frequency fell within the AC/DC component (50Hz for the UK), comparisons involving this sub-harmonic were not feasible. However there were no statistically significant differences between phase-locking of the stimulation frequency or its quarter-harmonic to cortical beta oscillations (p =0.1744, zval= -1.3583).

We also examined the synchrony of this 'supra-physiological' stimulation frequency with occipital oscillations, as we had for the 'physiological' stimulation frequency. This stimulation frequency was phase-locked to the gamma band of interest (p =0.0002, zval=3.6843) as well as occipital beta oscillations (p =0.0011, zval=3.2634, ranksum=148) to a significant degree compared to noise. When frontal and occipital effects were juxtaposed for trials in this stimulation frequency, the greatest effect was for a higher degree of phase synchrony with occipital alpha oscillations compared to frontal activity in this band (p =0.0032, zval=2.9467). The differences between frontal and occipital relationships of the stimulation to beta cortical oscillations bordered significance (p =0.0512, zval=1.9496).

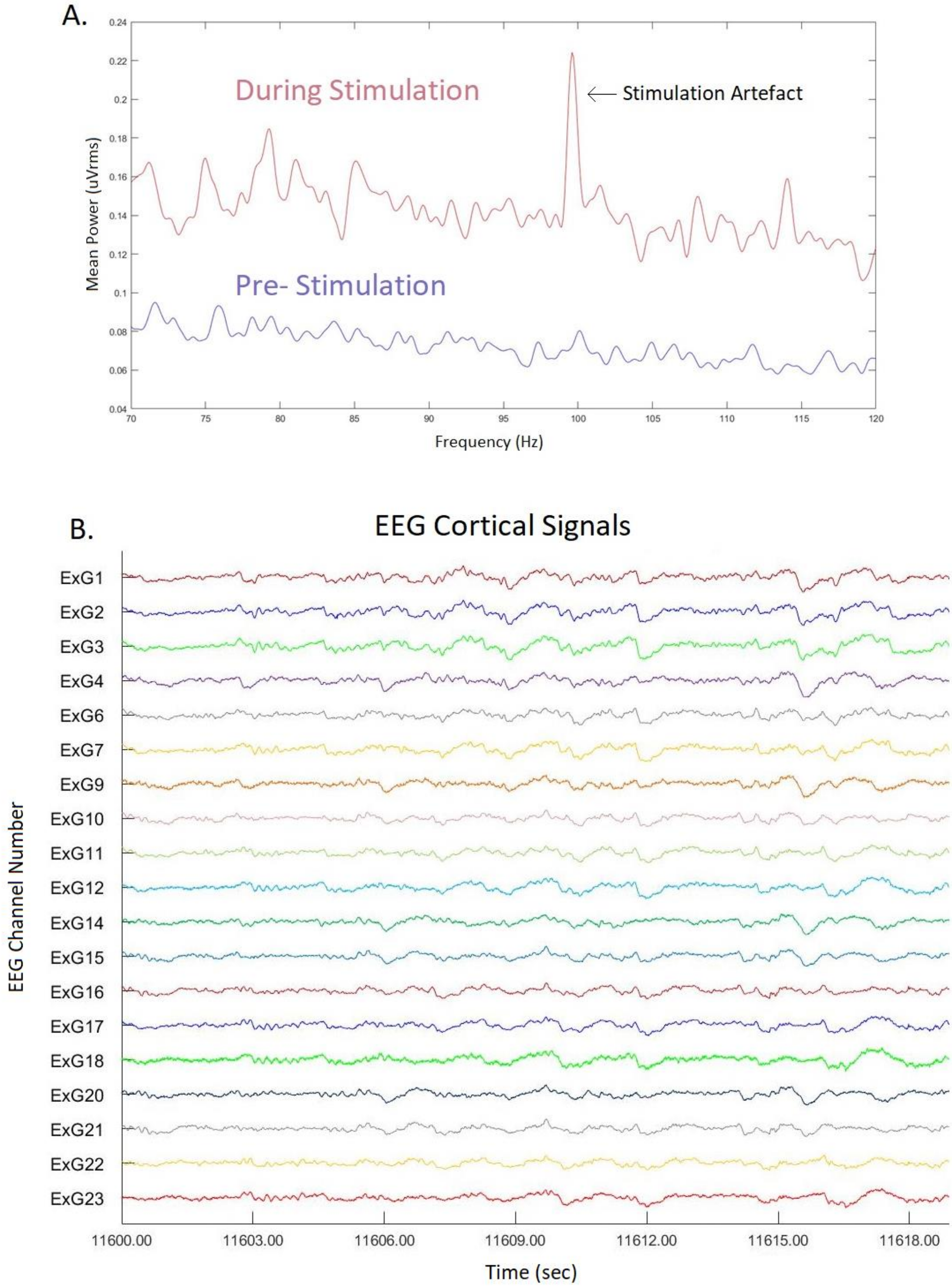


Supplementary Figure 7. Sample trial where, despite stimulation delivery, no artefact is present in EEG traces. *A. Linear plot of power spectra during stimulation delivery (unilateral at 100Hz, thresholded at 0.4mA for pulse width of 60µS). B. Due to the very low amplitudes, no stimulation artefact can be detected in the EEG traces (cortical signals). This trial had to be excluded from phase-locking analyses.*
